# MRI Intensity Standardization Evaluation Design for Head and Neck Quantitative Imaging Applications

**DOI:** 10.1101/2021.02.24.21252322

**Authors:** Kareem A. Wahid, Renjie He, Brigid A. McDonald, Brian M. Anderson, Travis Salzillo, Sam Mulder, Jarey Wang, Christina Setareh Sharafi, Lance A. McCoy, Mohamed A. Naser, Sara Ahmed, Keith L. Sanders, Abdallah S.R. Mohamed, Yao Ding, Jihong Wang, Kate Hutcheson, Stephen Y. Lai, Clifton D. Fuller, Lisanne V. van Dijk

**Affiliations:** Department of Radiation Oncology, University of Texas MD Anderson Cancer Center, Houston, TX.; Department of Imaging Physics, University of Texas MD Anderson Cancer Center, Houston, TX.; Department of Radiation Physics, University of Texas MD Anderson Cancer Center, Houston, TX.; Department of Head and Neck Surgery, University of Texas MD Anderson Cancer Center, Houston, TX.

## Abstract

**Background:** Conventional MRI poses unique challenges in quantitative analysis due to a lack of specific physical meaning for voxel intensity values. In recent years, intensity standardization methods to optimize MRI signal consistency have been developed to address this problem. However, the effects of standardization methods on the head and neck region have not been previously investigated.

**Purpose:** This study proposes a workflow based on healthy tissue region of interest (ROI) analysis to determine intensity consistency within a patient cohort. Through this workflow, we systematically evaluate different intensity standardization methods for T2-weighted MRI of the head and neck region.

**Methods:** Two image cohorts of five head and neck cancer patients, one with heterogeneous acquisition parameters (median age 59 years [range, 53-61]), and another with homogeneous acquisition parameters from a clinical trial (NCT03145077) (median age 61 years [range, 54-77]) were retrospectively analyzed. The standard deviation of cohort-level normalized mean intensity (SD NMI_c_), a metric of intensity consistency, was calculated across ROIs to determine the effect of five intensity standardization methods on T2-weighted images. For each cohort, the Friedman test with a subsequent post-hoc Bonferroni-corrected Wilcoxon signed-rank test was conducted to compare SD NMI_c_ among methods.

**Results:** Consistency (SD NMI_c_ across ROIs) between T2-weighted images is substantially more impaired in the cohort with heterogeneous acquisition parameters (0.28 ± 0.04) than in the cohort with homogeneous acquisition parameters (0.15 ± 0.05). Consequently, intensity standardization methods more significantly improve consistency in the cohort with heterogeneous acquisition parameters (corrected p < 0.005 for all methods compared to no standardization) than in the cohort with homogeneous acquisition parameters (corrected p > 0.05 for all methods compared to no standardization).

**Conclusions:** Our findings stress the importance of image acquisition parameter standardization, together with the need for testing intensity consistency before performing quantitative analysis of MRI.

## Introduction

Magnetic resonance imaging (MRI) is routinely used in clinical practice and has revolutionized how physicians evaluate disease pathology ^1^. Conventional “weighted” MRI acquisitions, where various acquisition parameters are modulated to generate T1-weighted (T1-w) or T2-weighted (T2-w) images, have become commonplace in physician assessment workflows. However, while conventional MRI acquisitions are currently useful for qualitative assessment of disease, advanced quantitative evaluation, such as through radiomics ^2^ or deep learning ^3^, is seemingly precluded by a fundamental problem: arbitrary voxel intensity. Unlike computed tomography (CT), where voxel intensities correspond to underlying inherent tissue characteristics, the absolute voxel intensities of MRI are a combination of tissue properties and hardware-specific settings ^4^, and thus do not have a specific physical meaning. Consequently, MRI voxel intensity can vary from scanner to scanner and even within the same scanner ^5^. A few important exceptions include images generated through various quantitative MRI acquisitions ^6^, such as diffusion-weighted MRI ^7^, dynamic contrast-enhanced MRI ^8^, or T1/T2 mappings ^9^, which are not routinely acquired in standard of care imaging. Unfortunately, intensity standardization (sometimes referred to as normalization or harmonization) is an often overlooked but assumed crucial pre-processing step in studies attempting a quantitative analysis of conventional MRI acquisitions (Fig. 1).

**Figure 1.**
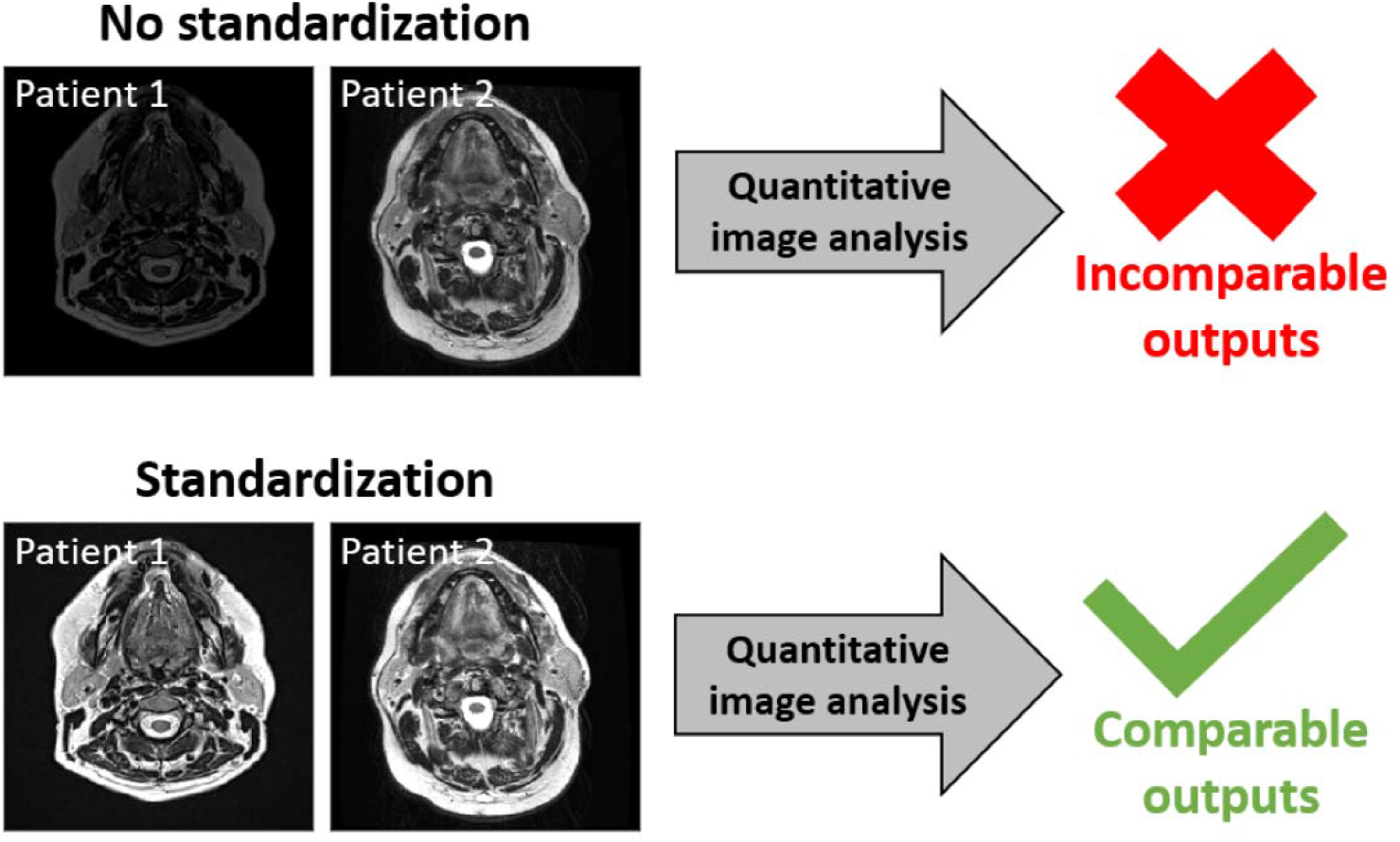
Intensity standardization may impact whether quantitative imaging analysis can be usefully conducted in conventional MRI of the head and neck region. Non-standardized T2-weighted images are displayed at a window width of 0 – 4000 arbitrary units while standardized images are displayed at a window width of −3 – 3 arbitrary units.

MRI scanning is often performed for head and neck cancer (HNC) patients as part of radiotherapy treatment. Weighted images, particularly T2-w images, are commonly acquired in the scanning protocol due to their excellent soft-tissue contrast in the complicated anatomic areas involved in HNC, and are thus of added value for tumor and healthy tissue delineation^10,11^. While many recent HNC studies have implemented quantitative analysis of conventional weighted MRI ^12–19^, relatively few explicitly incorporate intensity standardization in their methodological pipelines ^15–19^, with even fewer testing multiple methods ^19^. Moreover, while rigorous studies have tested MRI intensity standardization methods in various anatomical regions, chiefly in the brain ^5,20^, the head and neck region has yet to be systematically investigated. The head and neck region poses challenging problems when considering MRI intensity standardization, in particular when compared to relatively piecewise homogeneous regions like the brain. For instance, fields of view often vary across acquisitions, and the head and neck region is home to many tissue-tissue and tissue-air interfaces ^21^ that may increase difficulties when attempting to standardize intensities using the entire image. Therefore, there exists a pressing need to systematically investigate the effects of MRI intensity standardization methods in HNC.

To address these issues, this study sought to compare common intensity standardization methods for HNC T2-w images in a multi-institutional cohort with heterogeneous acquisition parameters and in a single-institutional cohort with homogeneous acquisition parameters.

## Methods

### Intensity Based ROI Evaluation

In line with the statistical principles of image normalization criteria introduced by Shinohara et al. ^5^, we assert that MRI intensities of the same tissue types should maintain similar distributions within and across patients, assuming no spatial inhomogeneities. Therefore, for a set of nonpathological regions of interest (ROI)s representing a corresponding set of tissues within a cohort of patients, an increasing quality of MRI intensity standardization should lead to a subsequent increase in the consistency of ROI intensity distributions (Fig. 2). It is important to note our aim is not to match distributions of the entire image since targets of quantitative image analysis (e.g., tumors or healthy tissues altered by radiotherapy such as the parotid glands) are expected to vary between patients.

**Figure 2.**
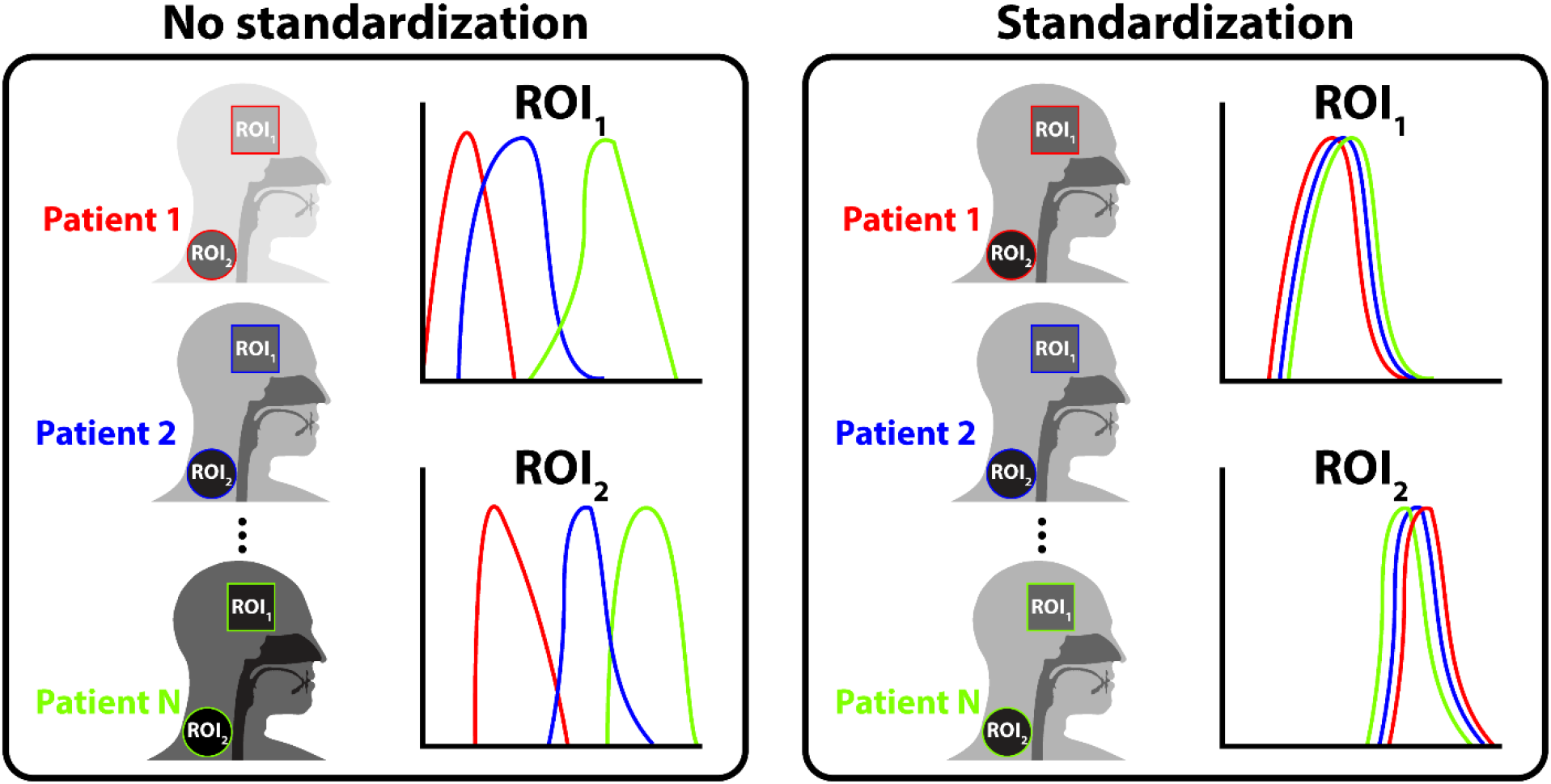
Intensity standardization qualitatively improves nonpathological region of interest (ROI) intensity distribution consistency at the cohort level. Nonpathological ROIs corresponding to healthy tissues with assumed phenotypic similarity are not expected to vary between patients; therefore, a proper intensity standardization aims to eliminate distributional differences between the same reference ROIs across patients.

Motivated by this goal of population-level analysis that relies on the consistency of replicable and biologically meaningful units within tissue types and across patients, we use a simple and interpretable metric of comparison to quantify ROI intensity histogram overlap which can be applied before or after an intensity standardization procedure in a given cohort. The steps to calculate this metric are given below and outlined visually in Figure 3:

1. Calculate the mean of a given ROI intensity distribution for a patient (μ*_p_*).
2. Divide μ*_p_* by the range of ROI intensity distributions for the entire cohort (*s*2*_c_* − *s*1*_c_*) thereby “localizing” the mean for that patient with respect to the entire distribution of values. The resulting value is the cohort-level normalized mean intensity (NMI_c_) for that ROI. s1_c_ and s2_c_ are set as the 2^nd^ and 98^th^ percentiles of ROI intensity distributions for the cohort, respectively, to remove any major outliers and were previously determined as appropriate from simulated data experiments (Supplementary Data S1).
3. Calculate NMI_c_ for each patient in the cohort and subsequently measure the “spread” of these values for the entire cohort, i.e., the standard deviation of NMI_c_ (SD NMI_c_) for that ROI.

**Figure 3.**
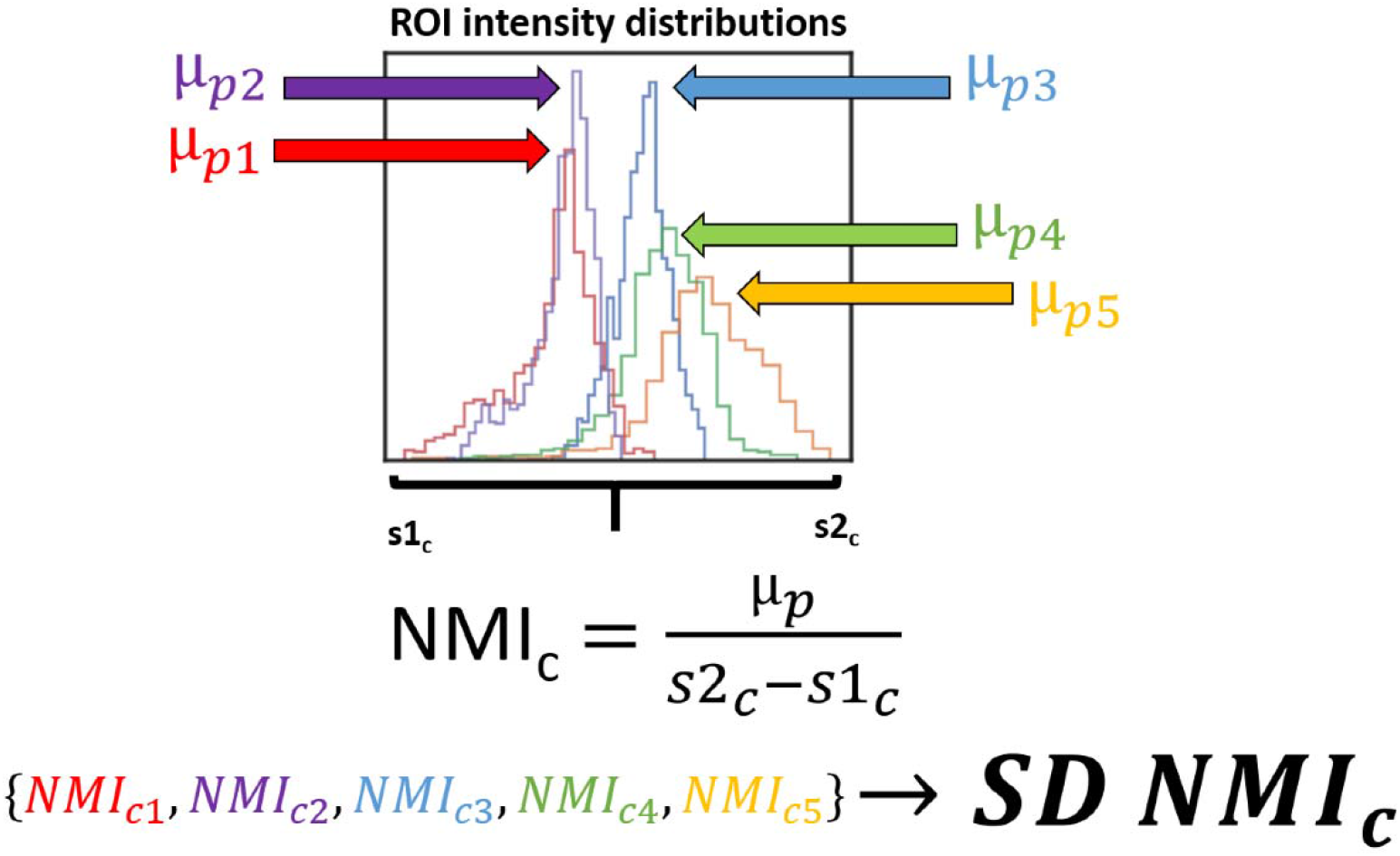
A newly derived metric, the standard deviation of cohort-level normalized mean intensity (SD NMI_c_), quantifies region of interest (ROI) intensity distribution overlap. By measuring the variability of distributional mean values on a normalized scale, SD NMI_c_ provides a scale-invariant cohort-level measure of intensity standardization consistency. SD NMI_c_ is determined by calculating the standard deviation of a set of cohort-level normalized mean intensity (NMI_c_) values. NMI_c_ values are calculated by dividing the mean of a given ROI intensity distribution for a patient (µ_p_) by the range of ROI intensity distributions for the entire cohort (s2_c_ − s1_c_).

SD NMI_c_ acts as a metric for characterizing the spread of centrality for a series of aggregated approximately Gaussian distributions. If we were to repeat this process for a given set of ROIs that are not anticipated to vary from patient-to-patient, we would expect that the cumulative SD NMI_c_ should remain as close to 0 as possible for an ideal intensity standardization method. SD NMI_c_, as defined here, is an adapted metric from Nyul et al. ^22^ where instead of normalizing with respect to a set of intensity values for a given patient, we normalize with respect to a set of intensity values for the entire cohort studied. SD NMI_c_ is scale-invariant, so it can robustly measure variation across different standardization methods for a given ROI without introducing bias based on the scale of standardization. We further compare SD NMI_c_ to other conventional metrics of distributional similarity in Supplementary Data S1.

### Patient Cohorts and Image Acquisitions

Two separate cohorts of five HNC patients diagnosed with oral or oropharyngeal cancer and with T2-w MRI images available before the start of radiotherapy were utilized for the analysis. The five patients from each cohort were randomly selected for this proof of concept study. The first cohort, labeled “heterogeneous” (HET), consisted of patients with images acquired at different institutions and is termed “heterogeneous” because of the variety of acquisition scanners and parameters used in image generation (Table 1). The second cohort, labeled “homogeneous” (HOM), consisted of patients from a single prospective clinical trial with the same imaging protocol (NCT03145077, PA16-0302) and is termed “homogeneous” because of the consistency in both scanner and acquisition parameters used in image generation (Table 2). Patients in the HOM cohort were immobilized with a head and neck thermoplastic mask during imaging. All images were retrospectively collected from the University of Texas MD Anderson Cancer Center clinical databases from the dates of May 2016 to October 2018 in agreement with an institutional review board and HIPAA approved protocol designed to collect data from patients with multiple imaging acquisitions (RCR03-0800). The protocol included a waiver of informed consent. The anonymized image sets analyzed during the current study are made publicly available online through Figshare (doi: 10.6084/m9.figshare.13525481, under embargo until formal manuscript acceptance in a scientific journal).

**Table 1.**
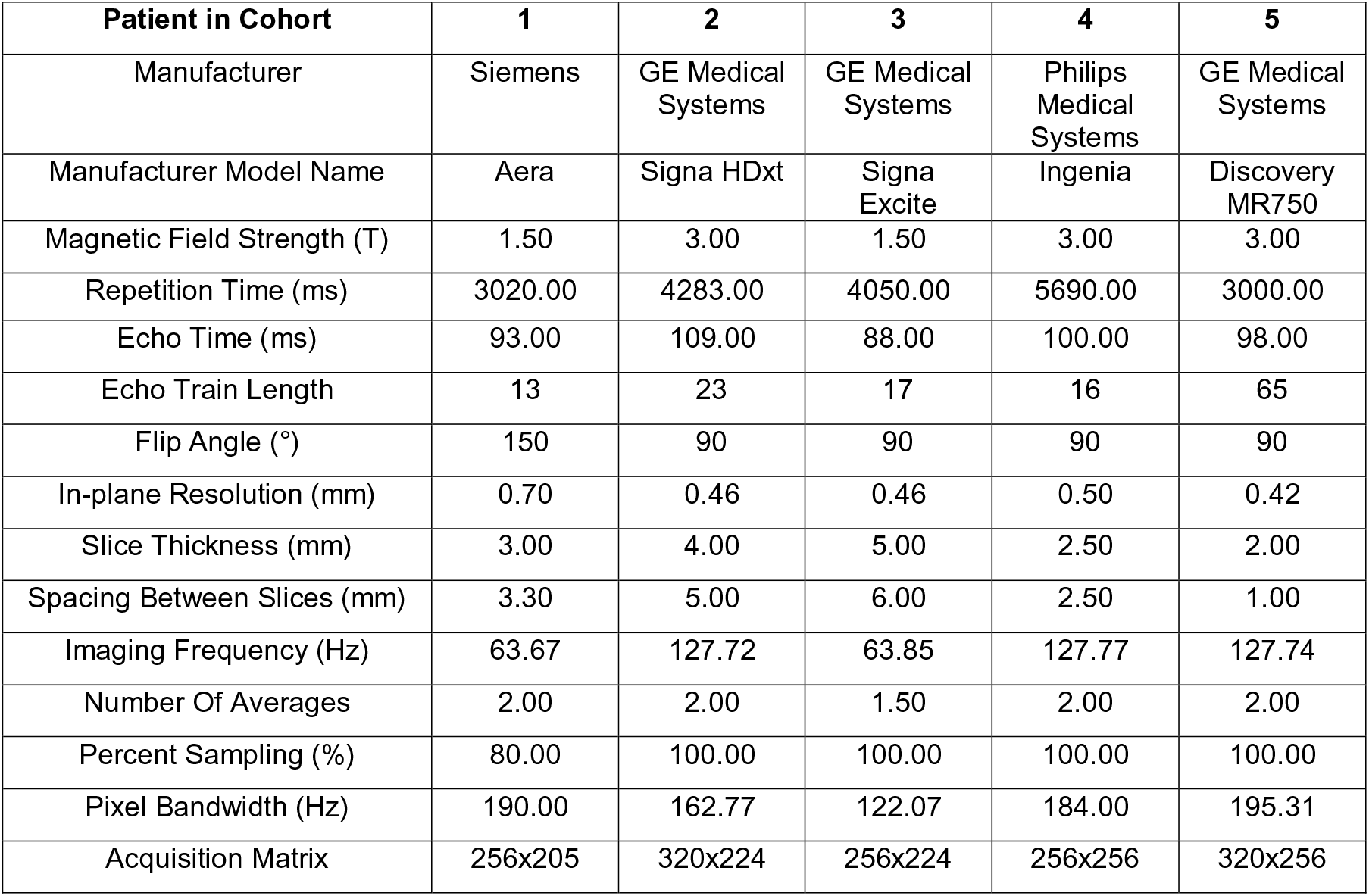
Heterogeneous (HET) cohort scanner characteristics. All five patients had images generated with different scanners and acquisition parameters.

**Table 2.** Homogeneous (HOM) cohort scanner characteristics. All five patients had images generated with the same scanner and acquisition parameters.

| Patient in Cohort | 1, 2, 3, 4, 5 |
| --- | --- |
| Manufacturer | Siemens |
| Manufacturer Model Name | Aera |
| Magnetic Field Strength (T) | 1.50 |
| Repetition Time (ms) | 4800.00 |
| Echo Time (ms) | 80.00 |
| Echo Train Length | 15 |
| Flip Angle (°) | 180 |
| In-plane Resolution (mm) | 0.50 |
| Slice Thickness (mm) | 2.00 |
| Spacing Between Slices (mm) | 2.00 |
| Imaging Frequency (Hz) | 63.67 |
| Number Of Averages | 1.00 |
| Percent Sampling (%) | 90.00 |
| Pixel Bandwidth (Hz) | 300.00 |
| Acquisition Matrix | 256x230 |

### Contours

For each image, ROIs from various healthy tissue types and anatomical locations were manually contoured in the same relative area for five slices by one observer (medical student) using Velocity AI v.3.0.1 (Atlanta, GA, USA) and verified by a physician expert (radiologist). The ROI names are shown in Table 3, and for representative ROI contours, we refer the reader to Figure S6.

**Table 3.** Regions of interest contoured for all patients. Structure abbreviations are shown in parenthesis and are used to refer to structures throughout the manuscript.

| Tissue category | Structures |
| --- | --- |
| Cerebrospinal fluid (CSF) | CSF inferior (CSF_inf), CSF middle (CSF_mid), CSF superior (CSF_sup) |
| Fat | Cheek Fat Left (Fat_L), Cheek Fat Right (Fat_R), Nape Fat Inferior (NapeFat_inf), Nape Fat Middle (NapeFat_mid), Nape Fat Superior (NapeFat_sup), Neck Fat (NeckFat) |
| Muscle | Masseter Left (Masseter_L), Masseter Right (Masseter_R), Rectus Capitus Posterior Major (RCPM) |
| Bone | Skull |
| Other | Cerebellum |

### Intensity Standardization Methods

We applied a variety of MRI intensity standardization methods to both cohorts. These methods were chosen because of their relative ubiquity in other studies and simple implementations. Details of the implementation of these methods are presented below. Method abbreviations are shown in parenthesis.

#### Unstandardized (*Original*)

No intensity standardization performed.

#### Rescaling (*MinMax*)

Standardizes the image by rescaling the range of values to [0,1].

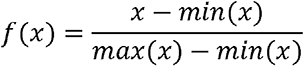

where *x* and *f(x)* are the original and standardized intensities, respectively, and *min(x)* and *max(x)* are the minimum and maximum image intensity values per patient, respectively. This method is widely used among deep learning MRI pipelines ^23–27^.

#### Z-score standardization using all voxels in image (*Z-All*)

Standardizes the image by centering it at a mean of 0 with standard deviation of 1. Standardization is based on all voxels in the image.

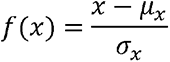

where *x* and *f(x)* are the original and standardized voxel intensities, respectively, and μ*_x_* and σ*_x_* are the mean and standard deviation of the image intensity values per patient, respectively. Currently the only intensity standardization method available before feature extraction in the popular open-source radiomic toolkit, PyRadiomics ^28^.

#### Z-score standardization using only voxels in an external mask (*Z-External*)

Z-score standardization as performed above but with respect to only voxels located in the external mask of head and neck region. The external mask is comprised of all non-background voxels from the top of the head down to between the sixth and seventh cervical vertebra and avoids lung spaces (Figure S4). Supplementary Data S2 demonstrates that using voxels above a pre-specified threshold acts as a proxy for the use of a head and neck external mask by eliminating signal nulls from lungs and outside the patient, which may be advantageous in cases where the generation of a mask is too time-consuming.

#### Cheek fat standardization (*Fat*)

Standardizes the image with respect to left and right cheek fat (healthy tissue). Method adapted from van Dijk et al. ^18^. Divides the intensity of each voxel by the mean of cheek fats and multiplies by an arbitrary scaling value of 350.

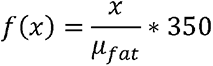

where *x* and *f(x)* are the original and standardized intensities, respectively, and μ*_fat_* is the mean of both cheek fat ROIs per patient. The utilization of healthy tissue for MRI standardization has been demonstrated in previous studies with mixed results ^29–31^. However, no previous studies have investigated the impact of healthy tissue standardization in the head and neck region.

#### Histogram standardization (*Nyul*)

Method adapted from Nyul and Udupa ^22^ with code implementation from Reinhold et al. ^32^. Utilizes images for all patients in a cohort to construct a standard histogram template through the determination of histogram parameters, then linearly maps the intensities of each image to the standard histogram. The standard histogram parameters in this implementation are defined as intensity percentiles at 1, 10, 20 … 90, and 99 percent. Only voxels within the head and neck external mask are used in the construction of the standard histogram.

### Displaying Standardized Images

All image manipulation and analyses were performed in Python v. 3.7.6 ^33^. Digital Imaging and Communications in Medicine (DICOM) images and radiotherapy structure files were converted to Python data structures using the DICOMRTTool package v.0.3.5 ^34^. While image dimensions for the HOM cohort had a fixed 512×512×120 array size, this was variable for the HET cohort. Image window width scaling for visualization was performed for each method by setting the maximum possible value at the 98^th^ percentile for the entire cohort. By employing this method, images could be visualized on a common scale per cohort with potential outliers removed for display purposes.

### Calculation of SD NMI_c_ and Significance Testing

For both cohorts, SD NMI_c_ was calculated for each intensity standardization method per ROI and visually compared on a heatmap. After applying a Shapiro-Wilk test ^35^ for normality, groups were found not to be normally distributed in the HET cohort (p < 0.05). Therefore, non-parametric tests were deemed appropriate for statistical analysis. For each cohort, the Friedman test ^36^, a non-parametric analog to the one-way repeat measures analysis of variance test, was conducted to compare SD NMI_c_ values among intensity standardization methods with standardization methods acting as within-subject factors. If the Friedman test was statistically significant, a subsequent post-hoc Wilcoxon signed-rank test with a Bonferroni correction ^37^ was performed with standardization methods acting as within-subject factors. For both the Friedman and Wilcoxon signed-rank tests, p-values < 0.05 were considered significant. Statistical analysis was performed in Python v. 3.7.6. Python code used to produce our analysis through Jupyter Notebooks are made publicly available through Github, which can be accessed at: https://github.com/kwahid/MRI_Intensity_Standardization.

## Results

To compare MRI intensity standardization methods in the head and neck region, we tested five methods in two distinct cohorts of HNC patients using SD NMI_c_ calculations in healthy tissues. We illustrate the overall workflow of our approach in Figure 4.

**Figure 4.**
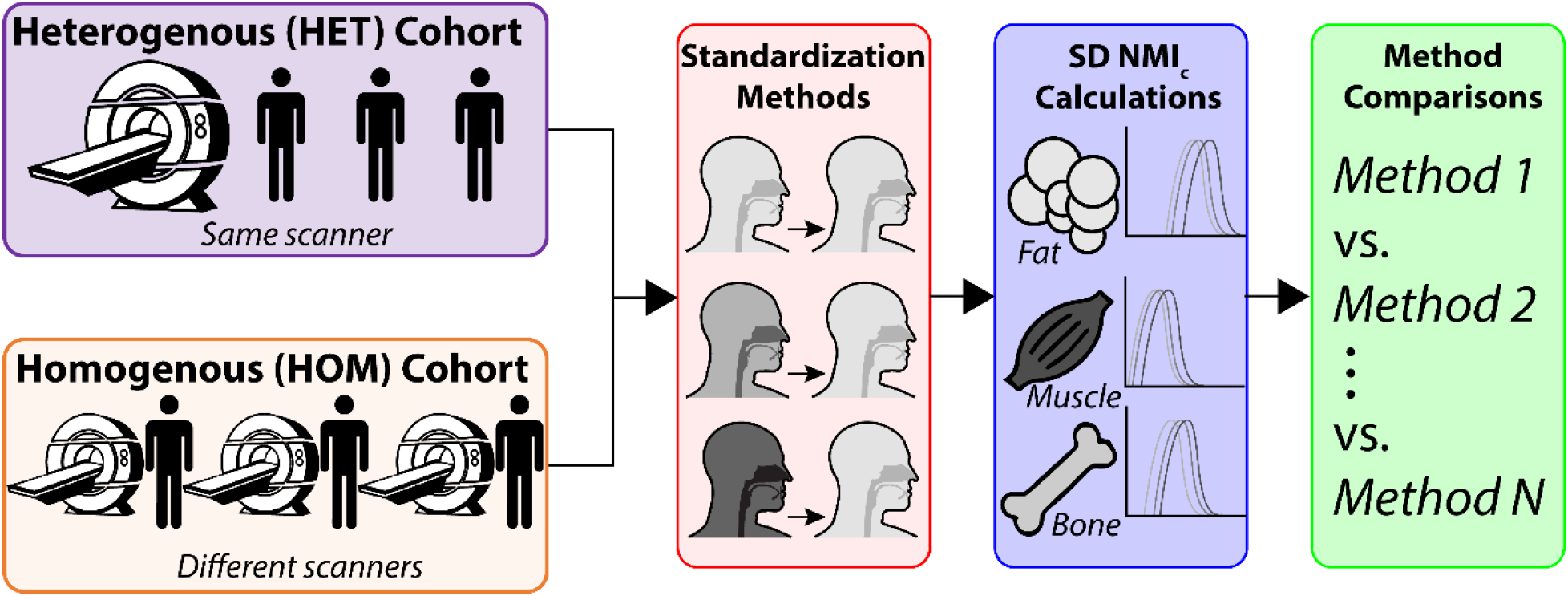
Steps in analysis workflow for this study. Intensity standardization methods are applied to two separate cohorts of HNC patients with either variable scanners/acquisition parameters (HET cohort) or consistent scanners/acquisition parameters (HOM cohort) and used to calculate standard deviation of cohort-level normalized mean intensity (SD NMI_c_) for distinct healthy tissues. Intensity standardization methods are subsequently compared in each cohort through significance testing.

### Demographic Characteristics

A total of five patients in the HET cohort (3 men, 2 women; median age of 59 years [range, 53-61]) and five patients in the HOM cohort (5 men, 0 women; median age of 61 years [range, 54-77]) were included in the analysis. The demographic characteristics for each cohort are summarized in Table 4.

**Table 4.** Patient characteristics for heterogeneous (HET) and homogeneous (HOM) cohorts. Unless otherwise indicated, data shown are number of patients with percentages in parenthesis.

| Characteristic | HET Cohort (n=5) | HOM Cohort (n=5) |
| --- | --- | --- |
| **Age (median, range)** | 59 (53-61) | 61 (54-77) |
| **Patient Sex** |  |  |
| Men | 3 (60) | 5 (100) |
| Women | 2 (40) | 0 (0) |
| **T Stage** |  |  |
| T1 | 4 (80) | 1 (20) |
| T2 | 1 (20) | 1 (20) |
| T3 | 0 (0) | 1 (20) |
| T4 | 0 (0) | 2 (40) |
| **N Stage** |  |  |
| N0 | 1 (20) | 0 (0) |
| N1 | 2 (40) | 3 (60) |
| N2 | 2 (40) | 2 (40) |
| **Primary Tumor Site** |  |  |
| Base of Tongue | 2 (40) | 2 (40) |
| Tonsil | 2 (40) | 3 (60) |
| Oral Cavity | 1 (20) | 0 (0) |

### Images and SD NMI_c_ Calculations

The unstandardized and standardized T2-w images for each of the patients from the HET and HOM cohorts are depicted in Figures 5a and 5c, respectively. Accordingly, heatmaps of the SD NMI_c_ values per ROI for each intensity standardization method for both cohorts are shown in Figures 5b and 5d. Additional representative T2-w images and ROI intensity distribution histograms for each patient in both cohorts are presented in Supplementary Data S3. For the HET cohort (Fig. 5b), mean SD NMI_c_ when averaged across all tissue sites adheres to the following SD NMI_c_ trends for each method (mean ± SD) from worst to best: *Original* (0.28 ± 0.04) > *MinMax* (0.21 ± 0.04) > *Z-All* (0.18 ± 0.04) > *Z-External* (0.16 ± 0.04) = *Fat* (0.16 ± 0.06) = *Nyul* (0.16 ± 0.04). For the HOM cohort (Fig. 5d), mean SD NMI_c_ when averaged across all tissue sites adheres to the following SD NMI_c_ trends for each method (mean ± SD) from worst to best: *Z-All* (0.16 ± 0.03) > *Original* (0.15 ± 0.05) > *Z-External* (0.14 ± 0.04) > *Fat* (0.13 ± 0.05) = *Nyul* (0.13 ± 0.04) > *MinMax* (0.12 ± 0.05).

**Figure 5.**
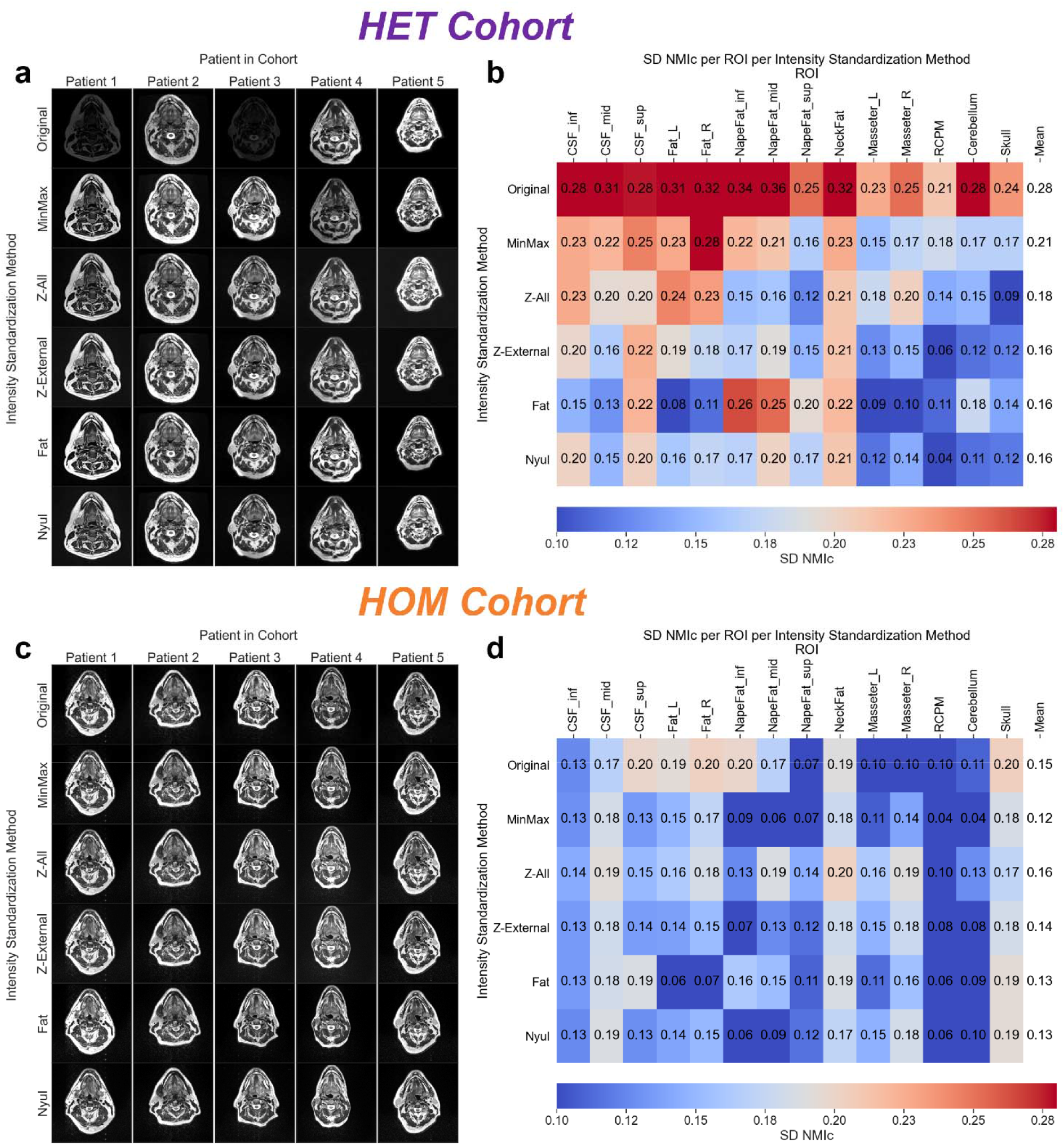
Intensity standardization comparisons for heterogeneous (HET) and homogeneous (HOM) cohorts. Single slice representation of T2-weighted images for each patient with respect to each intensity standardization method for HET **(a)** and HOM **(c)** cohorts. Images for each method in each cohort are displayed at the same window width. Standard deviation of cohort-level normalized mean intensity (SD NMI_c_) heatmaps of region of interest (ROI) with respect to intensity standardization method for HET **(b)** and HOM **(d)** cohorts. The resulting means across all ROIs for each method are shown in the rightmost column of heatmaps.

### Comparisons Between Intensity Standardization Methods

As depicted in Figure 6, the Friedman test showed that SD NMI_c_ values across all ROIs were significantly different for the intensity standardization methods in both cohorts (HET p<0.001, HOM p=0.001). On post-hoc analysis in the HET cohort (Fig. 6, above diagonal), significantly lower SD NMI_c_ values across all ROIs were found when comparing *Original* to *MinMax* (p<0.005), *Z-All* (p<0.005), *Z-External* (p<0.005), *Fat* (p<0.005), and *Nyul* (p<0.005) indicating these methods led to more consistent ROI intensities when compared to unstandardized images. Moreover, significantly higher SD NMI_c_ values were found when comparing *MinMax* to *Z-External* (p<0.005) and *Nyul* (p<0.005), indicating *MinMax* had less consistent ROI intensities when compared to the *Z-External* and *Nyul* standardization methods. In the HOM cohort (Fig. 6, below diagonal), no standardization methods had significantly different SD NMI_c_ values when compared to the unstandardized images, indicating a similar quality of ROI intensity consistency. Moreover, significantly higher SD NMI_c_ values were found when comparing *Z-All* to *MinMax* (p<0.01), *Z-External* (p<0.01), and *Nyul* (p<0.05), indicating *Z-All* had less consistent ROI intensities when compared to the *MinMax*, *Z-External*, and *Nyul* standardization methods.

**Figure 6.**
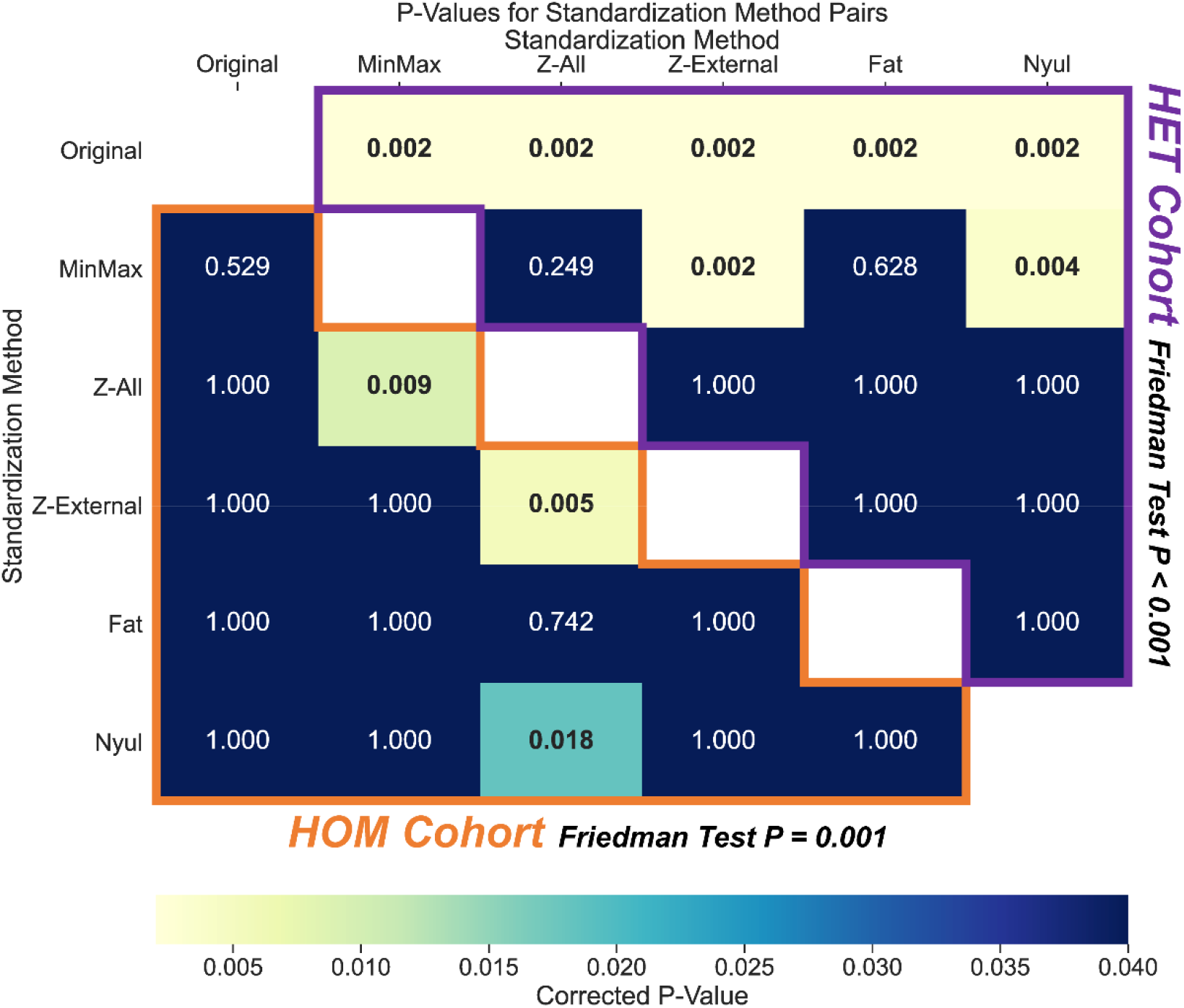
Statistical comparison matrix of standard deviation of cohort-level normalized mean intensity (SD NMI_c_) values between intensity standardization methods for heterogeneous (HET) and homogeneous (HOM) cohorts. Freidman test results are shown adjacent to the cohort titles. Each entry corresponds to a corrected p-value for a standardization method pair resulting from a Wilcoxon signed-rank test between SD NMI_c_ values for each healthy tissue ROI. Significant p-values (<0.05) are bolded in the heatmap. The HOM cohort results are outlined in orange below the white diagonal entries, while the HET cohort results are outlined in purple above the white diagonal entries.

We further investigated the effects of these various intensity standardization methods in other imaging modalities in the HOM cohort. For results concerning T1-w Dixon Water Suppressed MRI and radiotherapy planning CT scans (images, histograms, SD NMI_c_ heatmaps, and significance tests), we refer the reader to Supplementary Data S4 and S5, respectively. Moreover, to provide an example for a downstream quantitative imaging application, we perform additional analysis on T2-w images in both cohorts, comparing standardization methods with respect to radiomic feature categories in Supplementary Data S6.

## Discussion

In this study, we propose a workflow to test adequate consistency of standardized and unstandardized conventional MRI within a given HNC cohort for quantitative image applications. This is necessary, as unlike quantitative MRI techniques, which can be used to generate measurable maps of meaningful physical or chemical variables ^6^, no such natural image intensity mapping exists for conventional MRI acquisitions. The scale-invariant, and thus comparable, SD NMI_c_ metric was calculated to systematically investigate the effects of intensity standardization methods on T2-w images for two independent cohorts of HNC patients based on multiple healthy tissue ROI intensity consistency. Our analysis directly addresses the unmet need of intensity standardization in conventional MRI of the head and neck region.

Our results show that intensity standardization, when compared to no standardization, substantially affects T2-w MRI ROI intensity consistency in the HET cohort (Fig. 5a,b). Conversely, intensity standardization shows a minimal impact in the HOM cohort (Fig. 5c,d). Due to their vastly different scanner parameters, the vastly different variation in intensities discovered between the images from the HET cohort (Fig. 5a) was in line with our expectations. In contrast, for images based on large scale clinical trial data with the same scanner and acquisition parameters from the HOM cohort, the relatively minor variations in intensities between images (Fig. 5c) was better than expected. In a sense, the utilization of identical acquisition parameters in the HOM cohort seems to act as an inherent pre-processing intensity standardization. This also may indicate that the flexible head and neck MRI coil positioning was performed systematically and consistently in this cohort, as positioning of the coil with varying distances from the patient can result in different image intensities. Considering the ROIs individually in the HOM cohort, all intensity standardization methods showed considerably reduced SD NMI_c_ in Cheek Fat Left, Cheek Fat Right, and Nape Fat Inferior; however, intensity variation increased for muscular structures such as Masseter Left and Masseter Right (Fig. 5d). This discrepancy demonstrates the need to test standardization effects and careful consideration of the desired application space for the resulting intensity standardized images.

Generally, most intensity standardization methods performed similarly to each other in both cohorts, regardless of overall consistency improvement compared to unstandardized images (Fig. 5b,d and Fig. 6). A recent study by Carré et al. demonstrated similar results to our study in that various intensity standardization methods improved the consistency between images with heterogeneous acquisition parameters, but without identifying a specific superior standardization method ^38^. In the current study, paired significance testing (Fig. 6) revealed the standardization methods *Z-External* and *Nyul* performed relatively well in both the HET cohort (significantly better than *Original*) and the HOM cohort (significantly better than *Z-All*). While *Fat* had a low mean cumulative SD NMI_c_ equal to *Nyul* in both cohorts, paired significance testing revealed it did not perform as well as *Z-External* or *Nyul* in the HET and HOM cohorts. Moreover, the *MinMax* rescaling method performed best in the HOM cohort, while it performed worst in the HET cohort, suggesting this method is not robust. Interestingly, the only method that performed significantly worse than any others in the HOM cohort was *Z-All*, possibly secondary to misestimation of signal due to the large number of 0 elements influencing the standardization parameters in the HOM cohort.

This study focused on analyzing T2-w images for initial testing since they were readily available and are favored in head and neck imaging for their exquisite anatomic detail for a variety of ROIs ^39,40^. These results should be further investigated in images generated from other MRI sequences, but preliminary analysis shows similar trends for Dixon T1-w images in the HOM cohort (Supplementary Data S4). Interestingly, most intensity standardization methods significantly decreased ROI intensity consistency compared to unstandardized images for radiotherapy planning CT images (Supplementary Data S5), likely due to the already standardized Hounsfield unit scale being disrupted by the methods tested.

Upon visual inspection, it is difficult to discriminate between the various intensity standardization methods for either cohort (Fig. 5a,c). For the practicing clinician, small differences in standardization methods among conventional MRI acquisitions may not significantly affect image analysis, as the human eye can only distinguish a limited number of discretized gray levels ^41^. However, the subsequent downstream effects of standardization may be substantial when human eyes no longer become the driving force of analysis. For instance, Crombé et al. demonstrated marked differences in radiomic outcome prediction performance using different intensity standardization methods ^42^. We have performed a preliminary radiomic feature analysis comparing intensity standardization methods in Supplementary Data S6 which generally follows the trends highlighted in our study, but further work is needed to determine effects on HNC-specific outcome prediction tasks. Interestingly, Reinhold et al. showed that intensity standardization was crucial in deep learning MRI synthesis but did not observe large differences between standardization methods ^20^. Likewise, Jacobsen et al. showed that the specific method of standardization for deep learning segmentation is not as vital as other factors ^43^. These observations may indicate that deep learning is less sensitive to MRI intensity standardization than radiomic analysis, but further work should be performed to verify this.

One potential limitation of this proof of concept study is that we used a small number of patients (n=5) and distinct ROIs (n=13) for each cohort. Future work will include the addition of more patients and ROIs for each cohort to increase generalizability. Additionally, our ROIs are manually generated and, therefore, prone to inter/intra-observer variability ^44^. While ROIs were specifically chosen due to their consistency from patient to patient and assumed inherent biological similarity, auto-contouring workflows could reduce observer bias. Moreover, our analysis method may not apply to patients who have already received radiotherapy, as radiation could cause structural and functional changes in healthy tissue that may impact intensity profiles of various ROIs ^45^. This may be mitigated by selecting ROIs that are known not to change significantly with treatment. Finally, magnetic field inhomogeneities can affect these patients’ MRI intensity distributions, particularly for the HET cohort (Figure S18). Therefore, bias-field correction methodology may need to be explored to determine their effects on ROI intensity consistency.

To our knowledge, this is the first study systematically investigating the effects of intensity standardization in head and neck region imaging. Moreover, while many MRI intensity standardization studies implement test-retest data for individual patients to determine the effects of standardization ^29,38,46–48^, our analysis is unique by investigating the impact of standardization within a given cohort of patients. We propose this approach is more relevant to downstream cohort-level model construction often implemented in quantitative imaging studies. Finally, we have made our data and analysis tools available through open-source platforms to foster community reproducibility of our research. Our study is an essential first step towards widespread intensity standardization in conventional MRI for quantitative imaging of the head and neck region.

## Conclusions

Intensity standardization is key to improve the consistency of inherent tissue intensity values in conventional weighted MRI acquisitions between different patients and scanners in HNC images. In this study, five MRI intensity standardization methods were evaluated for T2-w HNC images at the cohort level with a proposed metric, SD NMI_c_, to quantify variation in normal tissue intensity values. Intensity standardization was found to be less pressing in a cohort with clinical trial regulated scanner and acquisition parameters than in a cohort for which these parameters were variable. We suggest that the implementation of homogeneous acquisition parameters in HNC can aid significantly in image intensity standardization. Moreover, post-acquisition intensity standardization methods can reduce inter-patient scan intensity variability and may positively affect downstream quantitative analyses. Specifically, after significance testing, histogram standardization and Z-score standardization using only voxels in an external mask performed relatively well for intensity consistency compared to other standardization methods in both tested cohorts. In summary, our study demonstrates the need for evaluating MRI intensity consistency before performing quantitative analysis and proposes a workflow to test this within a given HNC patient cohort.

## Data Availability

The anonymized image sets analyzed during the current study are made publicly available online through Figshare (doi: 10.6084/m9.figshare.13525481, under embargo until formal manuscript acceptance in scientific journal).

https://figshare.com/account/projects/95960/articles/13525481

## Acknowledgments

This work was supported by the Cancer Center Support Grant (P30-CA016672-44). K.A. Wahid and T. Salzillo are supported by training fellowships from The University of Texas Health Science Center at Houston Center for Clinical and Translational Sciences TL1 Program (TL1TR003169). R. He., A.S.R. Mohamed, K. Hutcheson, and S.Y. Lai are supported by a National Institutes of Health (NIH) National Institute of Dental and Craniofacial Research (NIDCR) Award (R01DE025248). B.A. McDonald receives research support from the NIH NIDCR (F31DE029093) and the Dr. John J. Kopchick Fellowship through MD Anderson UTHealth Graduate School of Biomedical Sciences. B.M. Anderson receives funding from the Society of Interventional Radiology Foundation Allied Scientist Grant and the Dr. John J Kopchick Fellowship. McCoy and K.L. Sanders are supported by NIH NIDCR Research Supplements to Promote Diversity in Health-Related Research (R01DE025248-S02 and R01DE028290-S01 respectively). M.A. Naser and S. Ahmed are supported by a NIH NIDCR Award (R01 DE028290-01). C.D. Fuller received funding from an NIH NIDCR Award (1R01DE025248-01/R56DE025248) and Academic-Industrial Partnership Award (R01 DE028290), the National Science Foundation (NSF), Division of Mathematical Sciences, Joint NIH/NSF Initiative on Quantitative Approaches to Biomedical Big Data (QuBBD) Grant (NSF 1557679), the NIH Big Data to Knowledge (BD2K) Program of the National Cancer Institute (NCI) Early Stage Development of Technologies in Biomedical Computing, Informatics, and Big Data Science Award (1R01CA214825), the NCI Early Phase Clinical Trials in Imaging and Image-Guided Interventions Program (1R01CA218148), the NIH/NCI Cancer Center Support Grant (CCSG) Pilot Research Program Award from the UT MD Anderson CCSG Radiation Oncology and Cancer Imaging Program (P30CA016672), the NIH/NCI Head and Neck Specialized Programs of Research Excellence (SPORE) Developmental Research Program Award (P50 CA097007) and the National Institute of Biomedical Imaging and Bioengineering (NIBIB) Research Education Program (R25EB025787). He has received direct industry grant support, speaking honoraria and travel funding from Elekta AB. L.V. van Dijk ceived/receives funding and salary support from the Dutch organization NWO ZonMw during the period of study execution via the Rubicon Individual career development grant.

## Supplementary Information

### Supplementary Data S1: Simulated Data Experiments

For the following experiments, we compare the standard deviation of cohort-level normalized mean intensity (SD NMI_c_) to the standard deviation of raw distribution means (SD_mean), and several commonly available python implementations of probability distribution similarity measures, including Jensen-Shannon distance (JSD), Wasserstein distance (Wasserstein), Kullback–Leibler divergence (KLD), and the Hellinger distance (Hellinger) ^1^. Where applicable, if multiple distributions are compared, we implement a pairwise mean calculation of the similarity measures.

#### Experiment 1: SD NMI_c_ Metric Fail Cases and Empirically Guided Selection of s1_c_ and s2_c_ Values

To remove bias from our analysis, we select the s1_c_ and s2_c_ values at percentage thresholds so outliers do not overly influence SD NMI_c_. However, selecting appropriate values for s1_c_ and s2_c_ is critical since unexpected behavior may occur if a large enough fraction of SD NMI_c_ values are calculated past these thresholds. In these sets of experiments, we seek to determine at what point the SD NMI_c_ metric no longer returns reliable values for comparing distributions and subsequently select reasonable values for s1_c_ and s2_c_ based on simulated data experiments and empirical observations of real data. First, we determine maximally physiologically plausible values to move a certain percentage of outliers away from the other distributions in the cohort. To determine this, for all regions of interest (ROI)s in the heterogeneous (HET) cohort, we measure the maximum spread between the means of the distributions furthest from each other (leftmost vs. rightmost) in the unstandardized images. These maximum values are obtained for Cheek Fat Left at a range of 3199.13 arbitrary intensity units. Using this maximum range, we perform a simulation experiment on 1000 arbitrary distributions where we progressively increase the number of distributions at this range, setting s1_c_ and s2_c_ to 5 and 95, respectively. We observe that for outlier values from 1 to 5%, the SD NMI_c_ value increases at a rate not indicative of the underlying difference in distributions, being heavily influenced by outliers until moving past 5%. This is compared with the Wasserstein distance and KLD (Hellinger is equivalent to Wasserstein for visualization purposes, and JSD is unable to characterize the spread of non-overlapping distributions at such a distance), which have expected monotonic behavior (Figure S1). We suggest that the selection of s1_c_ and s2_c_ should reflect an observed physiological maximum percentage of values outside the means of the furthest distributions. Therefore, we again utilize unstandardized images from the HET cohort to determine a reasonable set of values for s1_c_ and s2_c_. Towards this end, we measure the percentage of values less than the highest patient mean, which we find corresponds to Nape Fat Inferior at 96.7%. We conservatively increase this value to 98% and set it as s2_c,_ and correspondingly set s1_c_ to 2%. We suggest the use of these values to avoid spurious behavior in future studies.

**Figure S1.**
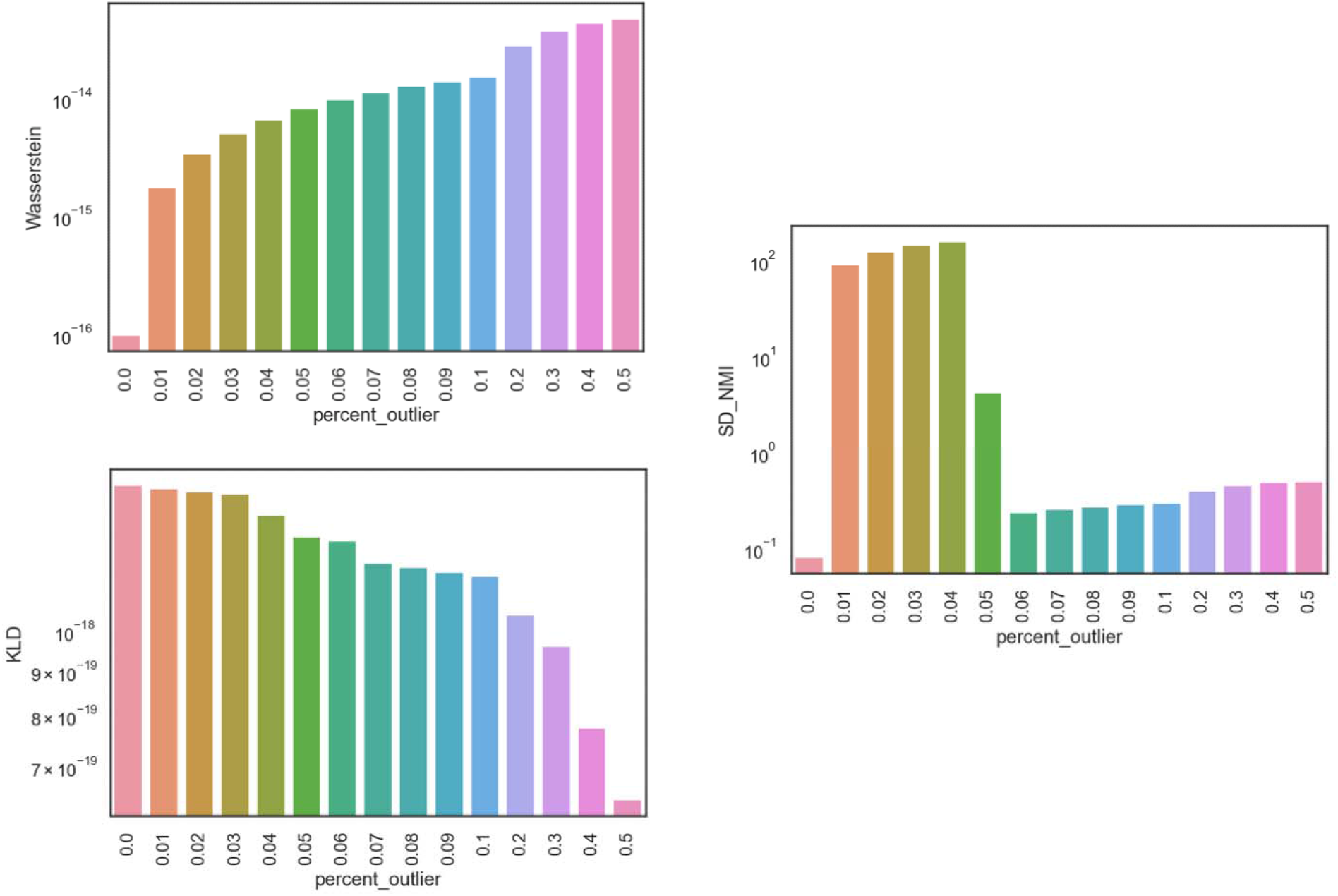
As the percentage of outliers increases above a threshold, the standard deviation of cohort-level normalized mean intensity (SD NMI_c_) metric (right) behaves erratically when compared to Wasserstein distance (Wasserstein) (top left) or Kullback–Leibler divergence (KLD) (bottom left).

#### Experiment 2: Properties of SD NMI_c_ and Other Measures of Distributional Similarity After Application of Scaling Factor

In this set of experiments, we aim to determine if SD NMI_c_ is invariant to arbitrary scaling of distributional data, an important property for comparing cohorts of ROIs before and after standardization. This is particularly relevant for determining an adequate metric for MRI intensity scaling quality, as one could erroneously interpret a change in standardization quality by applying arbitrary scaling factors that alter a chosen measure. We apply scaling factors of 2, 3, and 4 to two arbitrary distributions, calculate similarity measures, and plot normalized values (Fig. S2). Subsequently, the only measure invariant to scale is SD NMI_c_, reinforcing its appropriateness as a metric for our analysis.

**Figure S2.**
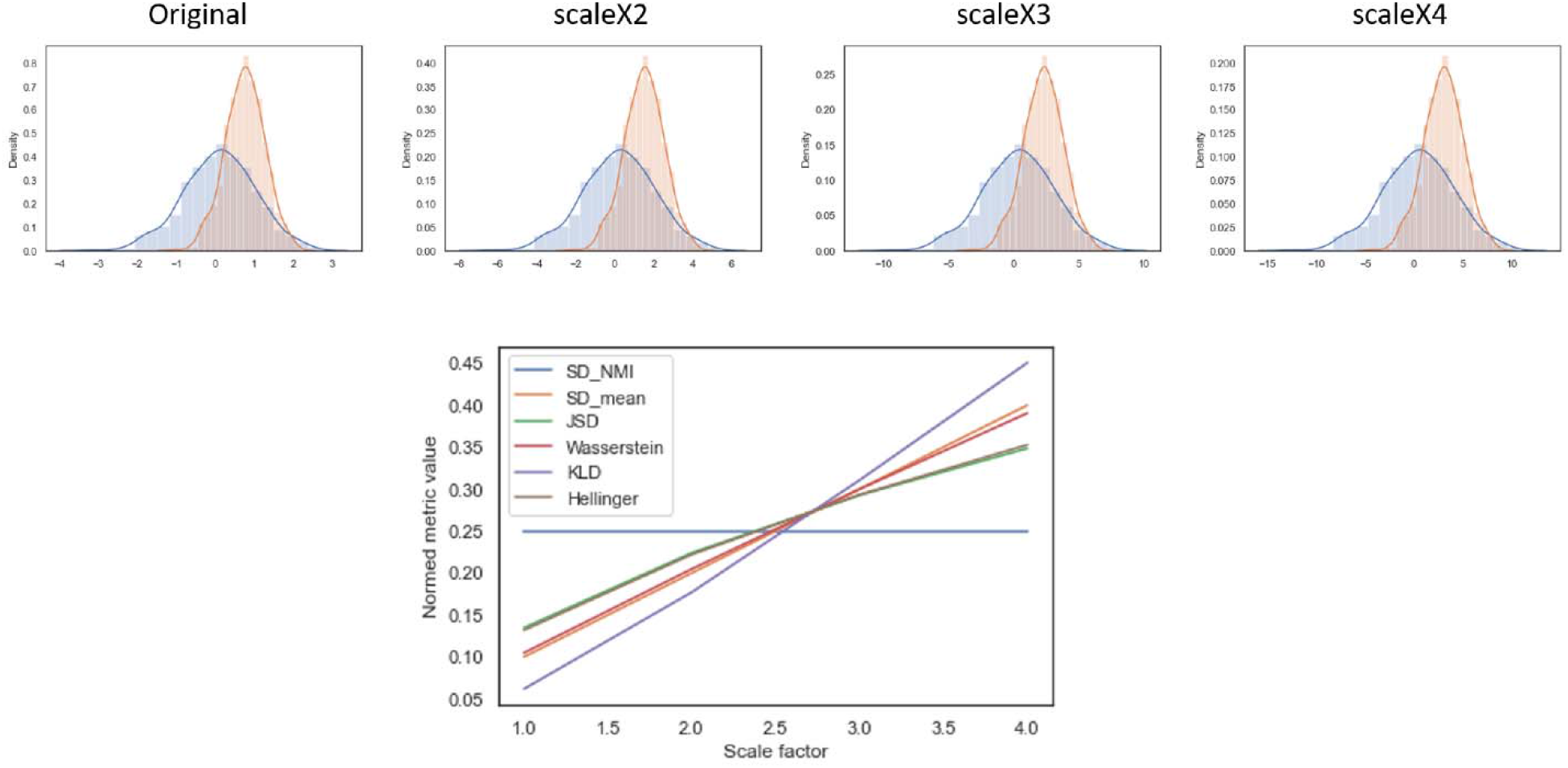
An increasing scale factor does not affect standard deviation of cohort-level normalized mean intensity (SD NMI_c_) (blue line) while it affects other measures of distributional similarity (other colored lines).

#### Experiment 3: Correlative Characterization of SD NMI_c_ With Respect to Other Measures of Distributional Similarity

In these sets of experiments, we attempt to determine the impact of histogram (distributional) parameters on similarity measures and the relationships between similarity measures. To accomplish this, we bootstrap 1000 random variations of histogram parameters (mean, standard deviation, skew, kurtosis) and determine the impact of these parameters (mean pairwise difference of mean, standard deviation, skew, kurtosis) on calculated similarity measures through correlative analysis. In this case, we generate synthetic data for five distributions as an analog to the five patients per cohort we analyze in our main study. All parameters are between −1 and 1, except for the standard deviation, which is between 0 and 1. As shown in Figure S3, SD NMI_c_ is most heavily impacted by the standard deviations of a set of distributions followed by the means of a set of distributions, which logically emerges from its definition. SD NMI_c_ is not highly correlated to any of the conventional similarity measures (JSD, Wasserstein, KLD, Hellinger). A clear trend exists between SD NMI_c_ and SD_mean because of their similar approach to calculating the variance of means in a cohort. Additionally, prominent correlative trends exist between conventional measures of distributional similarity (JSD, Wasserstein, KLD, Hellinger), which are expected due to their similar statistical properties ^2^.

**Figure S3.**
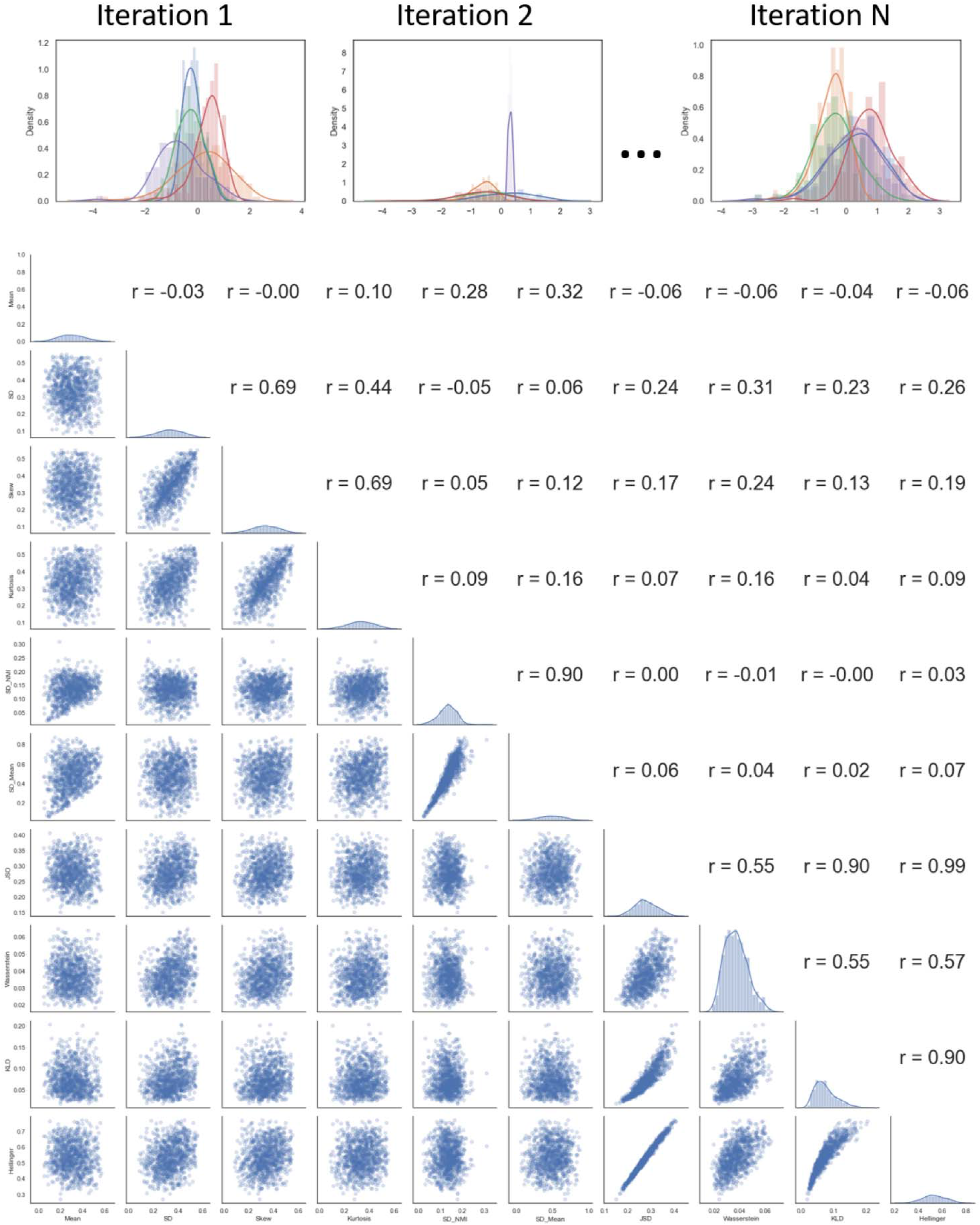
Simulation experiments of repeated iterations of distributional parameter resampling (top graphs) reveal relationships between different measures of distributional similarity (SD NMI_c_, SD_mean, JSD, Wasserstein, KLD, Hellinger) and distributional parameters (mean pairwise difference of mean, standard deviation (SD), skew, kurtosis). Standard deviation of cohort-level normalized mean intensity = SD NMIc; standard deviation of raw distribution means = SD_mean; Jensen-Shannon distance = JSD; Wasserstein distance = Wasserstein; Kullback– Leibler divergence = KLD; Hellinger distance = Hellinger.

### Supplementary Data S2: Image-Mask Visualizations

#### External Mask Example

**Figure S4.**
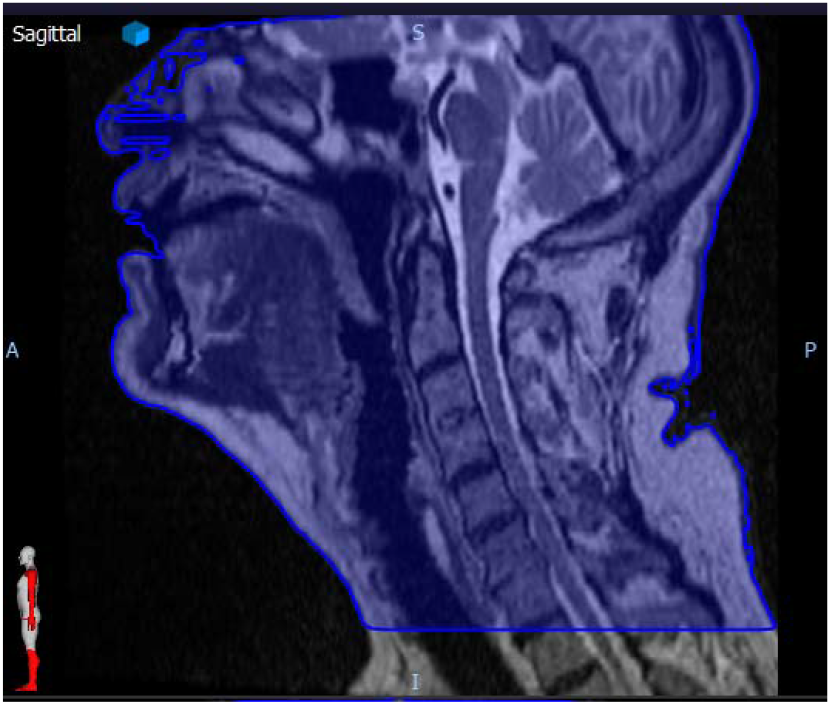
Example of an external mask (blue outline) for a head and neck cancer patient in the homogeneous (HOM) cohort.

#### External Mask Approximation

We analyzed intra- and inter-patient variation in whole image signal intensity of patients from the homogeneous (HOM) cohort by measuring the mean voxel intensity per slice on an individual basis (slice 1, slice 2, slice 3, …). An example of the slice by slice directionality moving inferiorly to superiorly is shown in Figure S5 (top images). Three types of variations on the analysis of slice statistics were performed: *1. All voxels examined for calculations, 2. Only voxels with values >30 examined, 3. Only voxels contained within the external mask examined*. Graphs of mean voxel intensity per individual slice for all techniques per patient are displayed in Figure S5 (bottom graph). We demonstrate that utilizing voxels with values >30 closely approximates the variation in selecting an external mask. Since the maximum voxel value in the HOM cohort is on the order of 500 arbitrary intensity units, we recommend using a voxel cutoff of 6% (i.e., only include voxels > 6% of maximum intensity value) to generalize to arbitrary cohorts.

**Figure S5.**
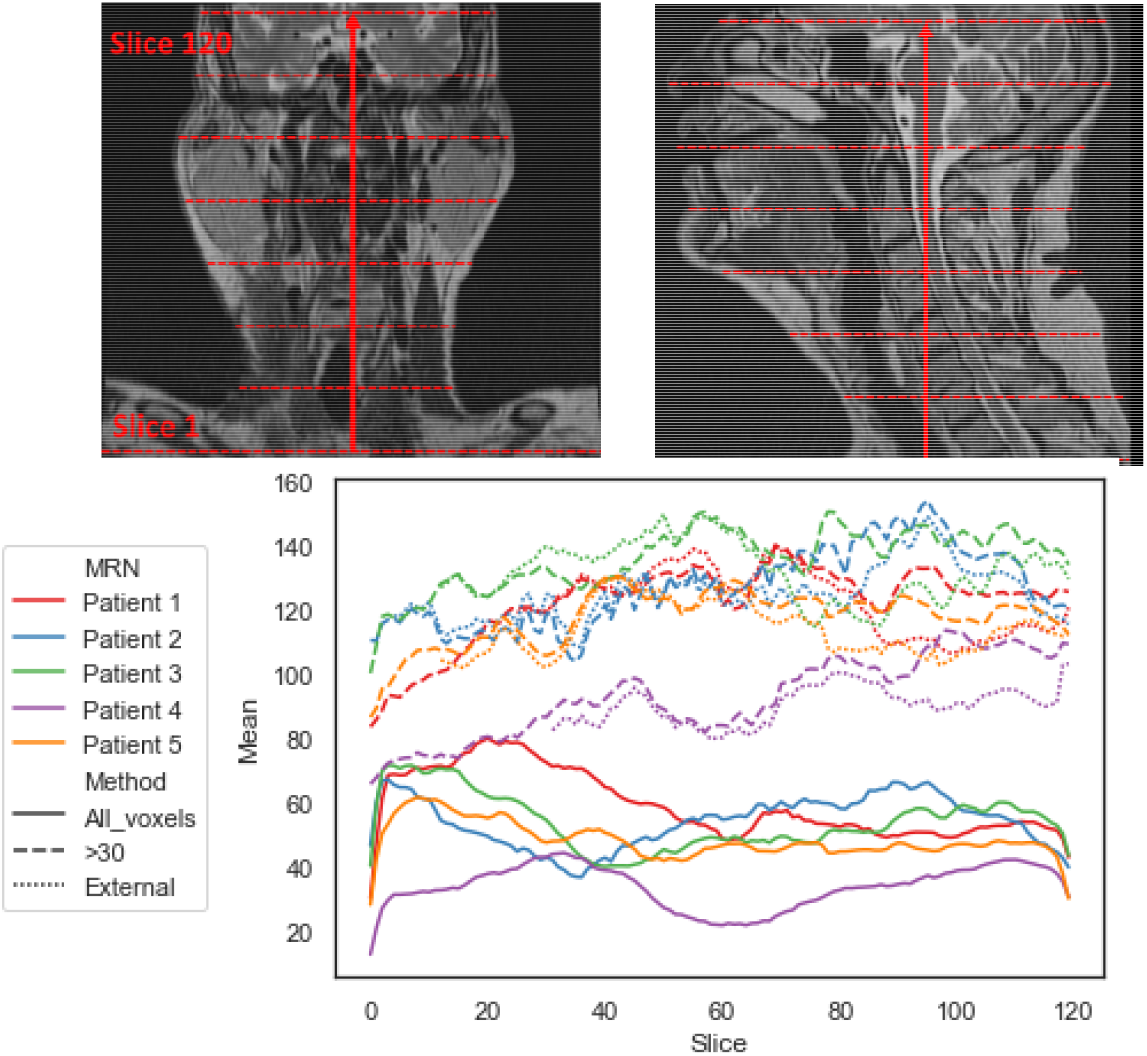
Mean voxel intensity per individual slice per patient for T2-weighted images in the homogeneous (HOM) cohort. Slices were analyzed in a bottom-top manner. Utilizing voxels with intensity values >30 closely approximates the variation in selecting an external mask (dashed lines compared to dotted lines).

#### Example of ROIs for a HNC Patient in the HOM Cohort

**Figure S6.**
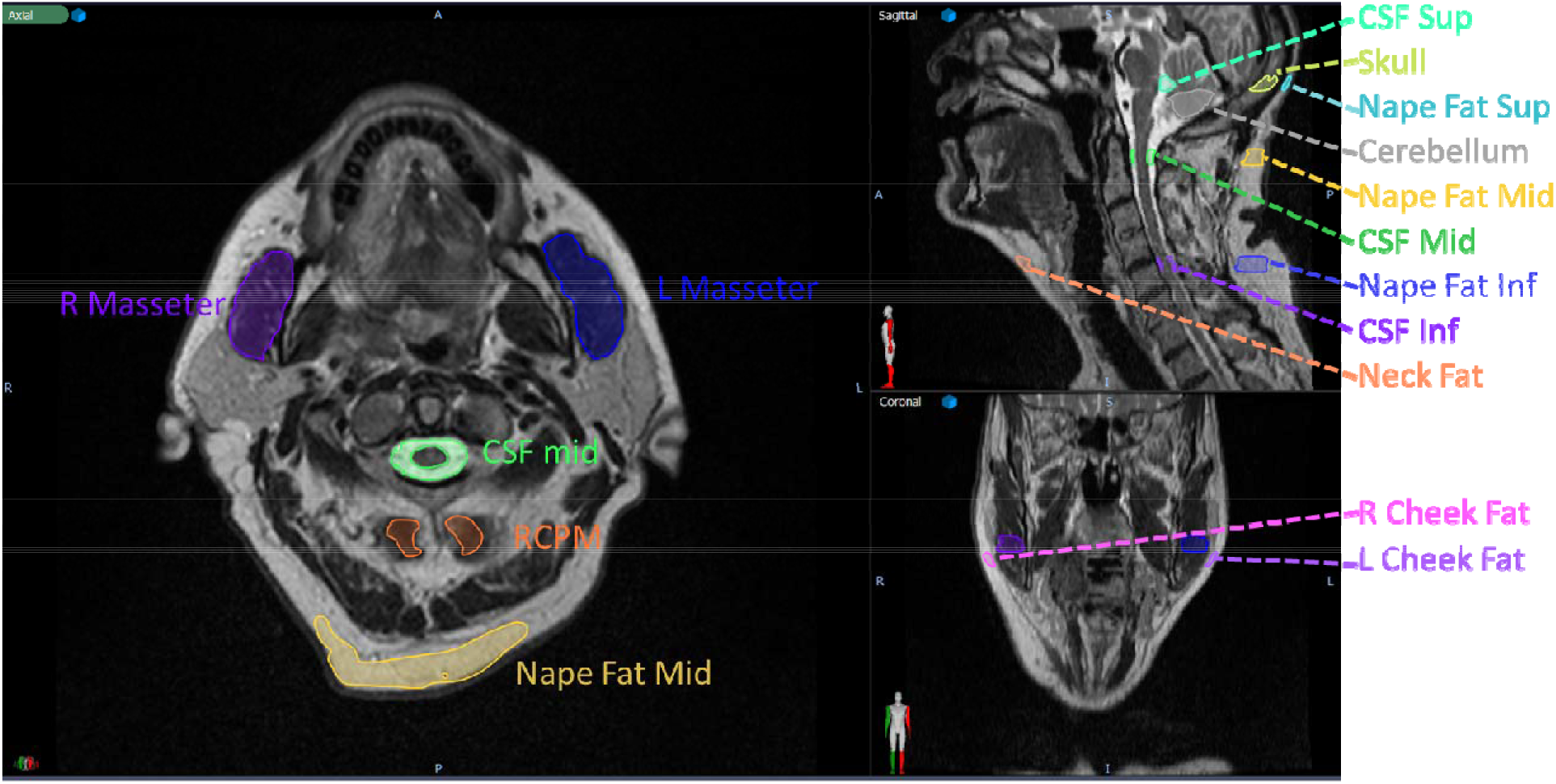
Various regions of interest consisting of tissue types from different anatomical locations were contoured for all patients. All regions of interest were contoured for 5 slices in the same relative area for all patients. T2-weighted image shown is an example from the homogeneous (HOM) cohort.

### Supplementary Data S3: Complete T2-w Data

#### HET Cohort Histograms

**Figure S7.**
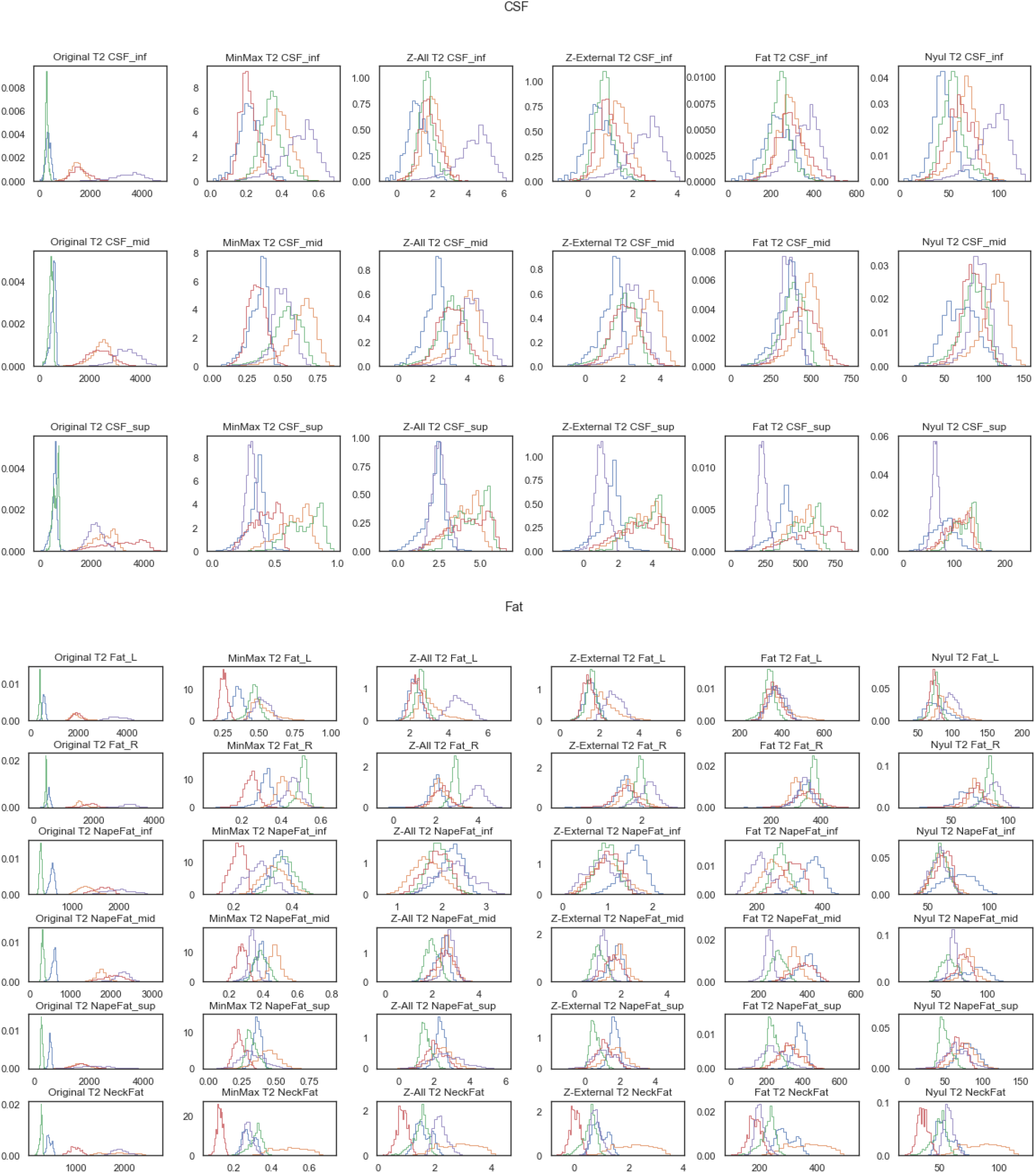
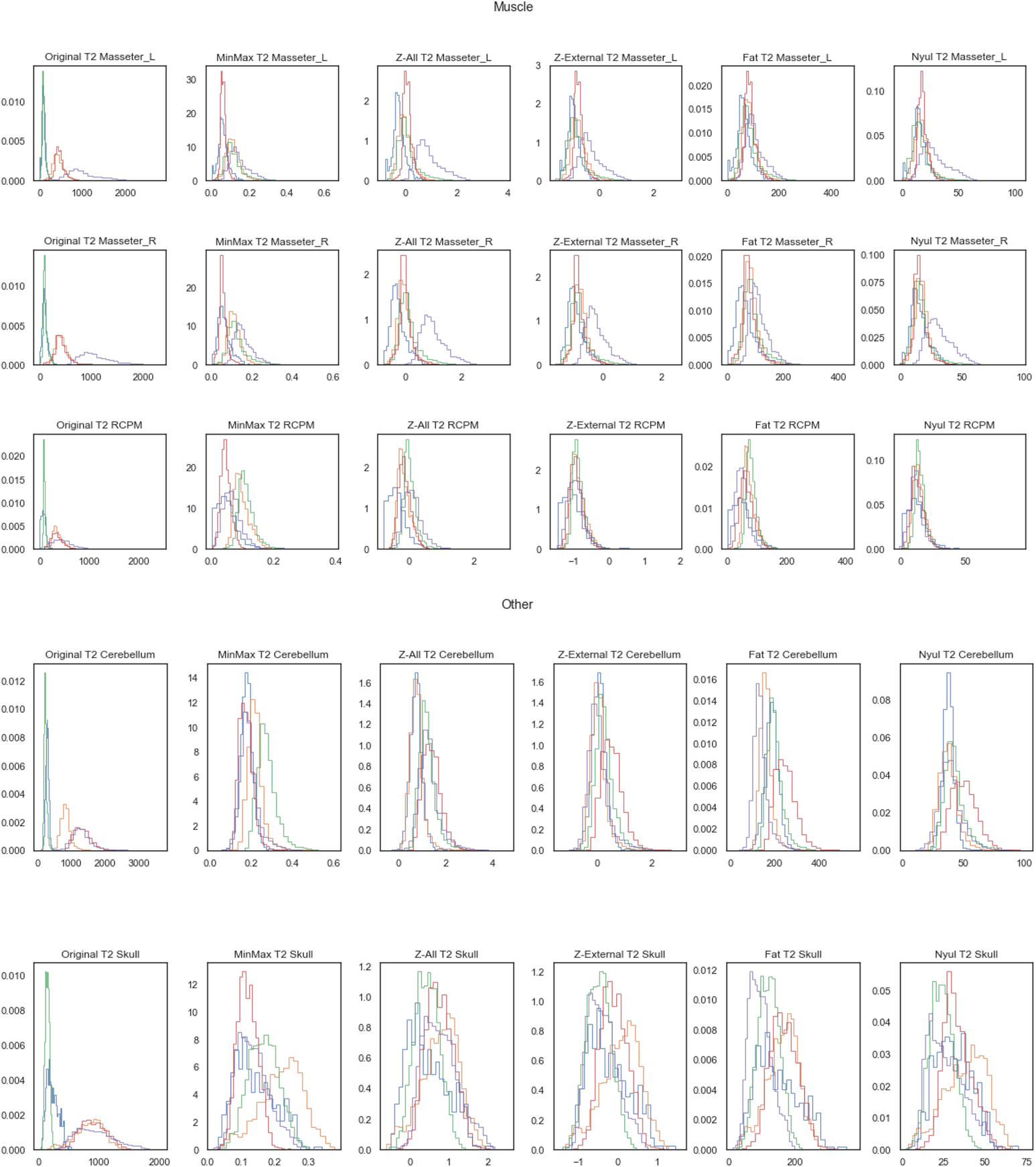
Distributions of region of interest voxel intensities for all patients in the heterogeneous (HET) cohort for T2-weighted images. Different colored histograms correspond to different patients in cohort. Columns correspond to methods while rows correspond to regions of interest. First, second, third, and fourth sets of plots correspond to CSF, Fat, Muscle, and Other categories, respectfully.

#### HET Cohort Images

**Figure S8.**
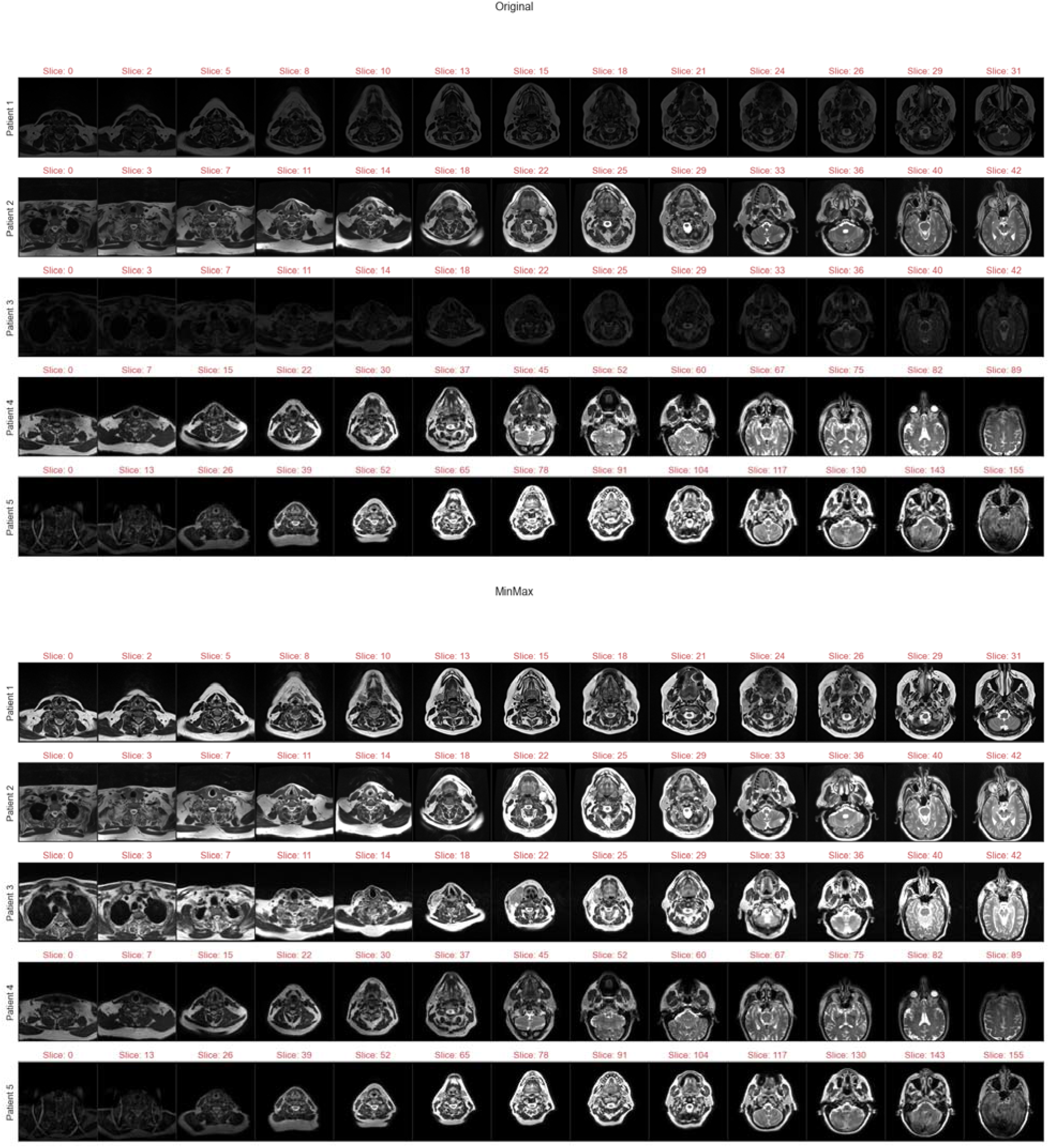
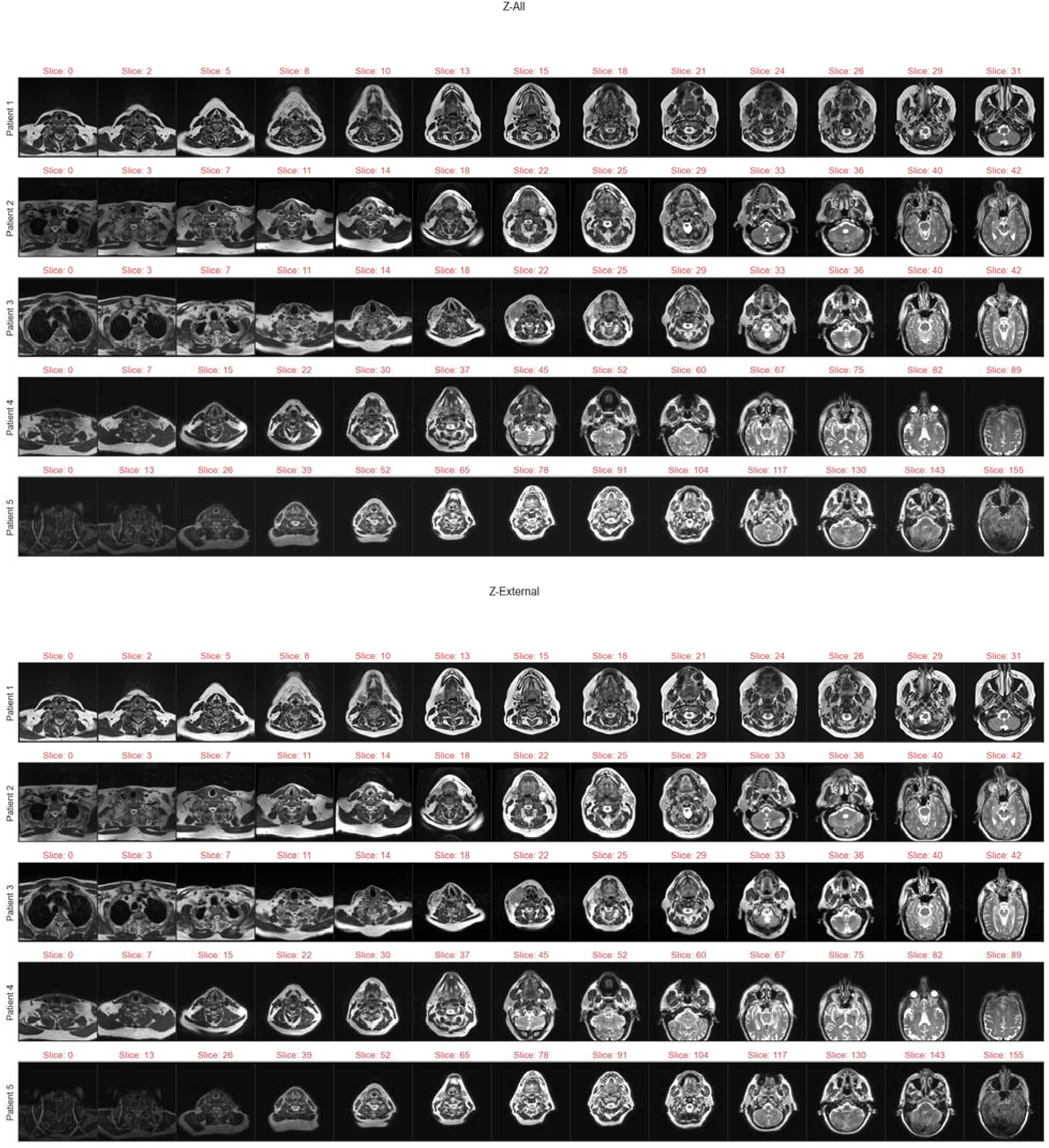
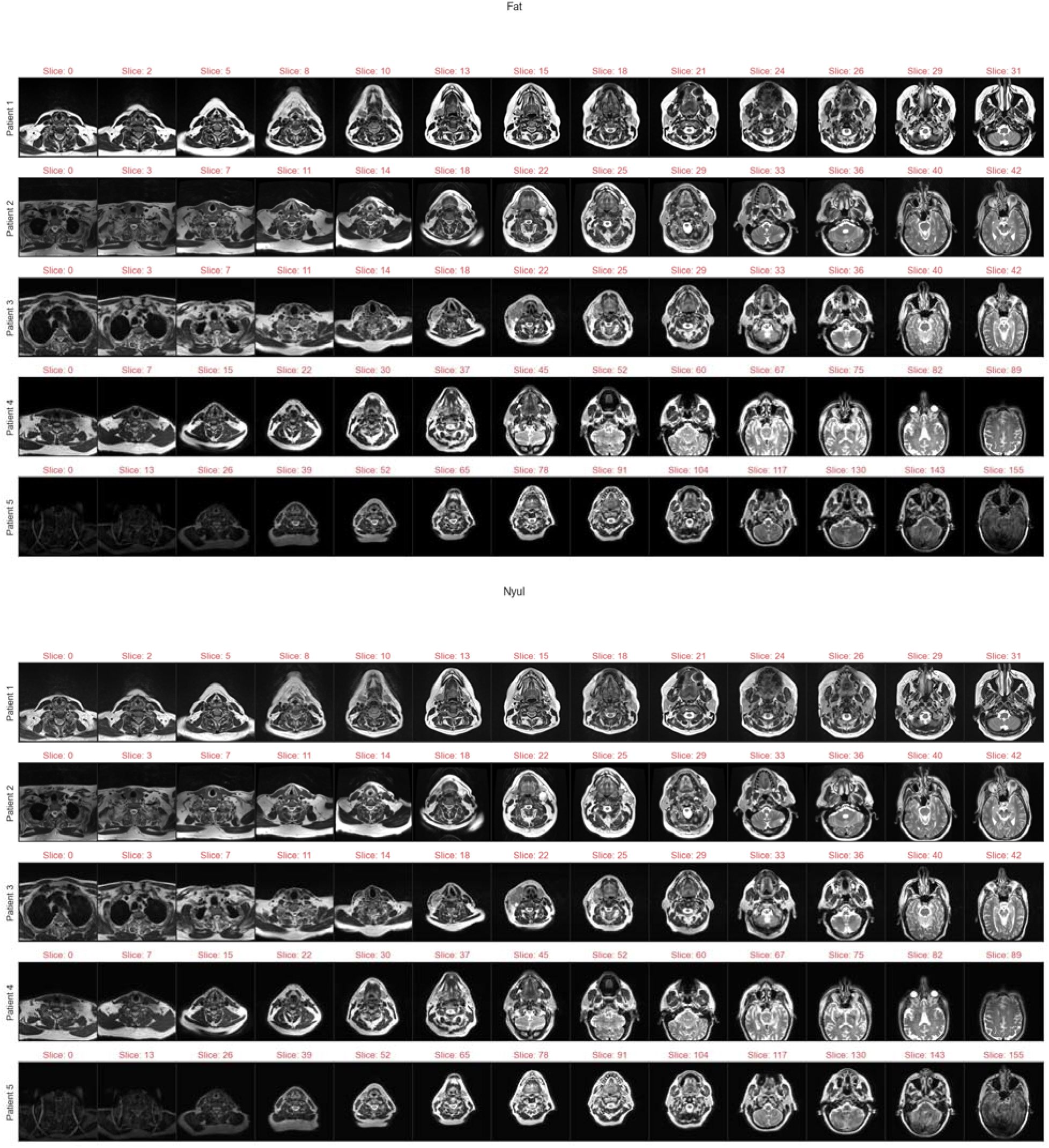
Example T2-weighted images for each patient in heterogeneous (HET) cohort. Images are at intervals of 1/13 total image slices to visualize full field of view for each patient. First, second, third, fourth, fifth, and sixth sets of plots correspond to Original, MinMax, Z-All, Z-External, Fat, and Nyul standardization methods, respectively.

#### HOM Cohort Histograms

**Figure S9.**
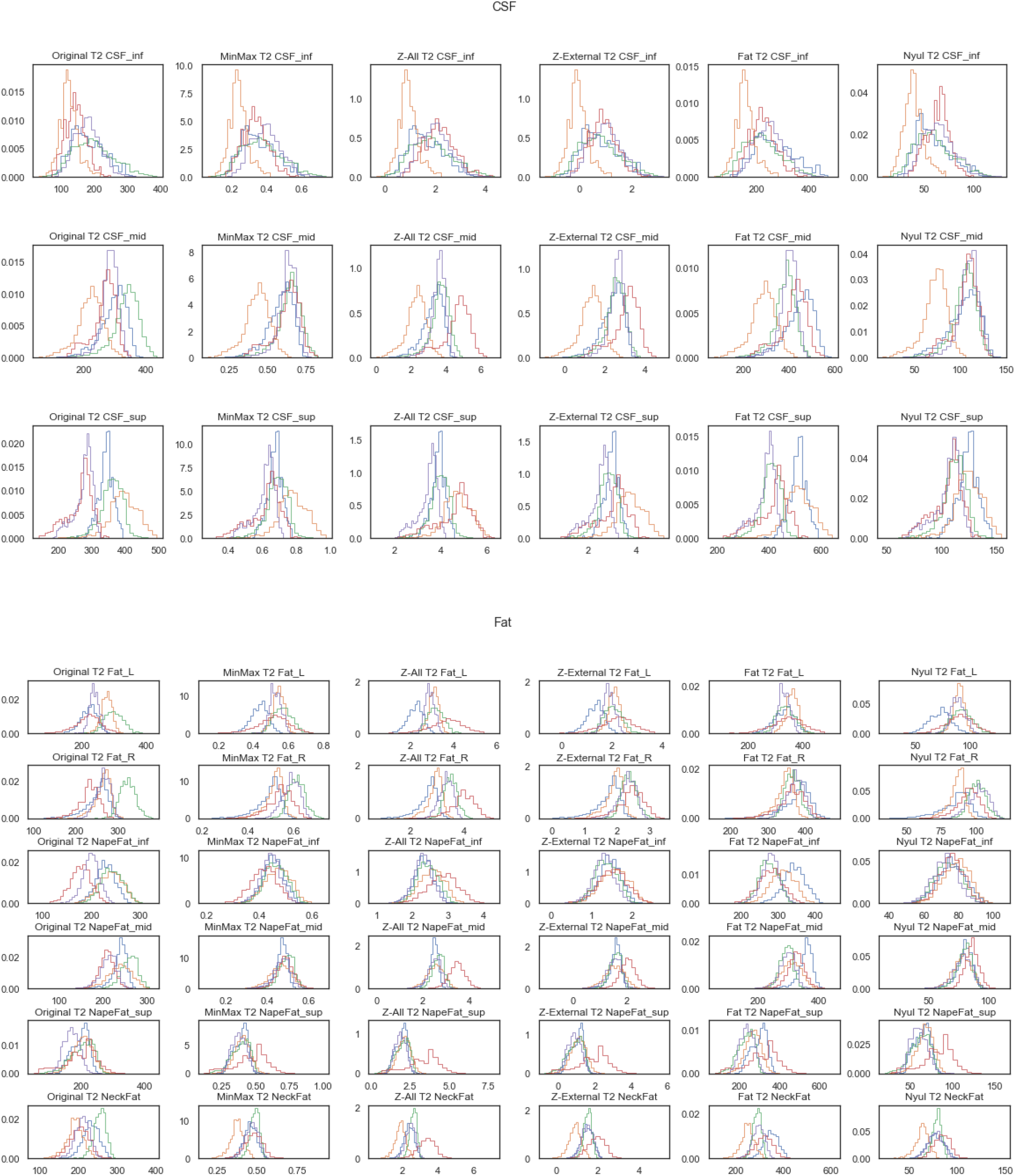
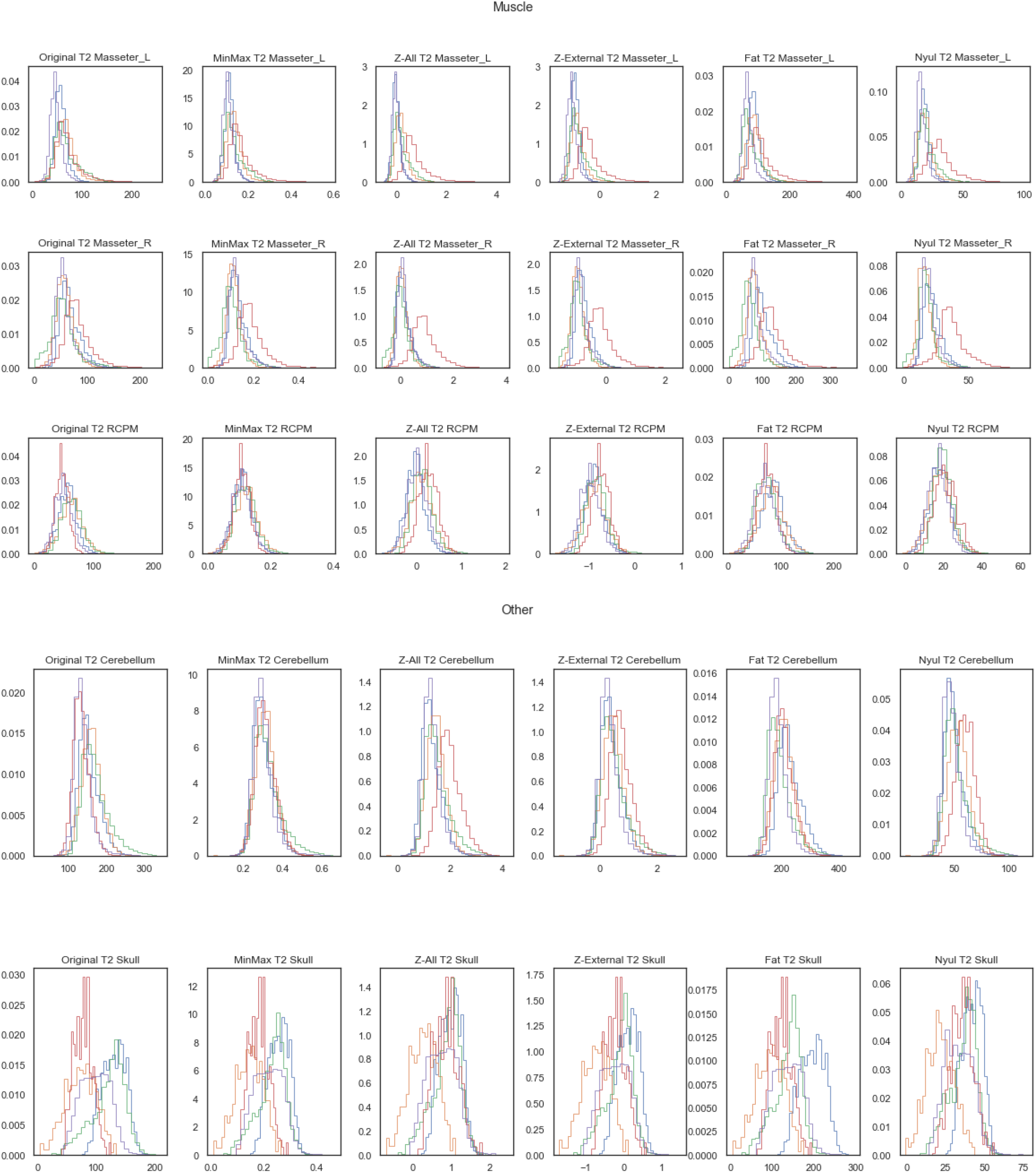
Distributions of region of interest voxel intensities for all patients in the homogeneous (HOM) cohort for T2-weighted images. Different colored histograms correspond to different patients in cohort. Columns correspond to methods while rows correspond to regions of interest. First, second, third, and fourth sets of plots correspond to CSF, Fat, Muscle, and Other categories, respectfully.

#### HOM Cohort Images

**Figure S10.**
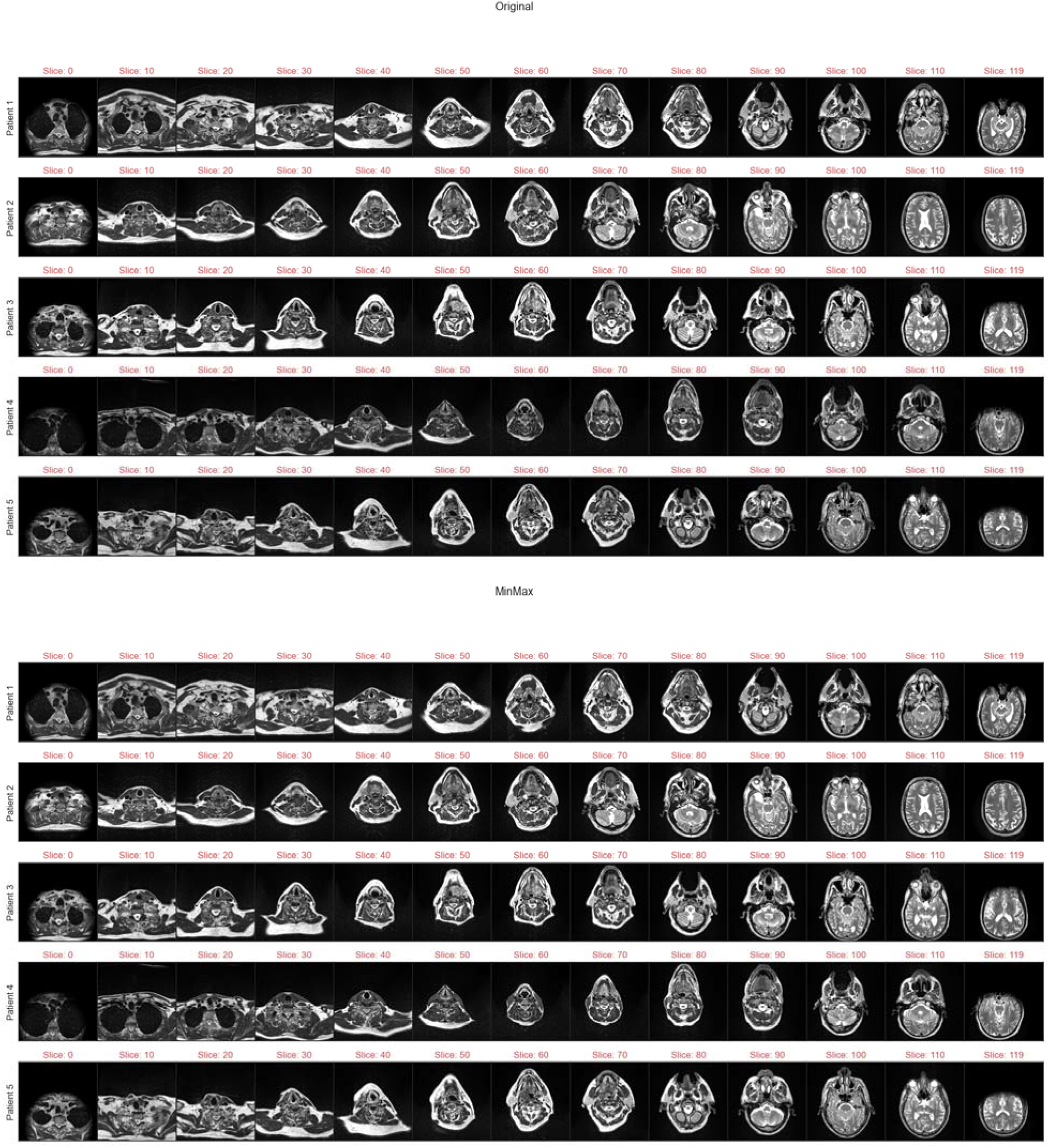
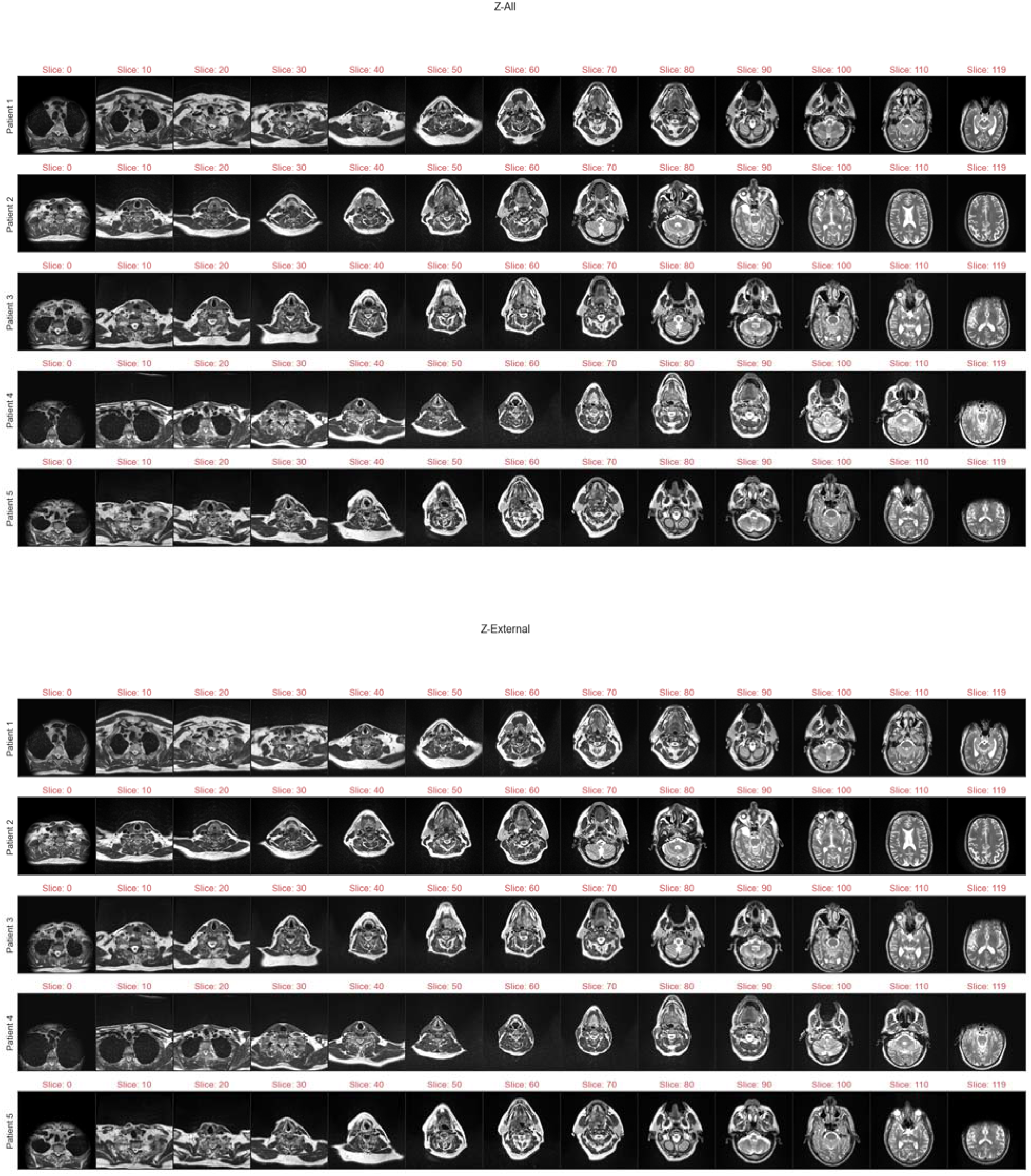
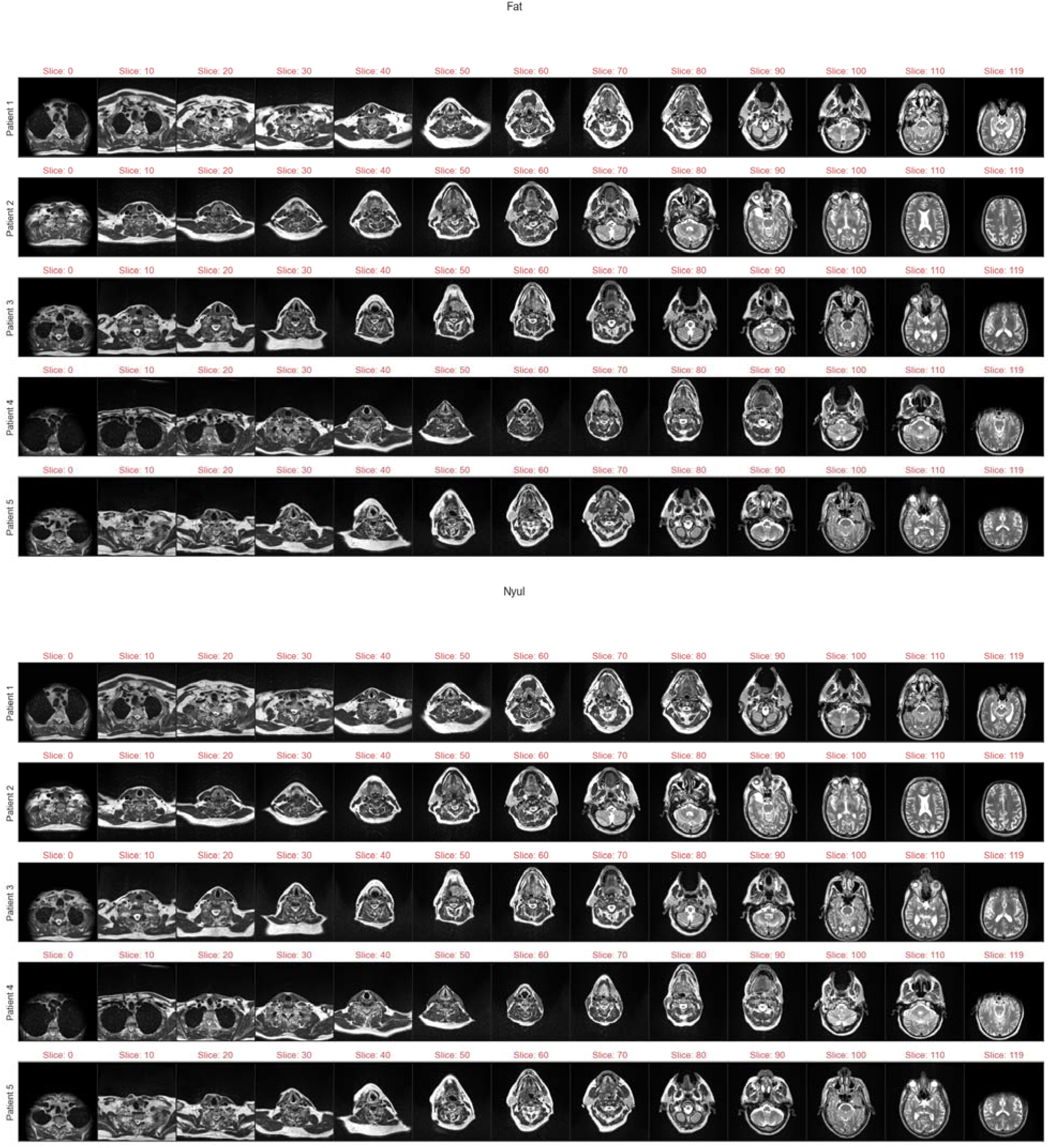
Example T2-weighted images for each patient in homogeneous (HOM) cohort. Images are at intervals of 1/13 total image slices to visualize full field of view for each patient. First, second, third, fourth, fifth, and sixth sets of plots correspond to Original, MinMax, Z-All, Z-External, Fat, and Nyul standardization methods, respectively.

### Supplementary Data S4: HOM Cohort T1-w Dixon Water Suppressed MRI Data

#### HOM Cohort T1-w Dixon Water Suppressed Scanning Parameters

**Table S1.** Homogeneous (HOM) cohort scanner characteristics for Dixon T1-w Water Suppressed images. All five patients had the same scanner/acquisition parameters.

| Patient in cohort | 1, 2, 3, 4, 5 |
| --- | --- |
| Manufacturer | Siemens |
| Manufacturer Model Name | Aera |
| Magnetic Field Strength (T) | 1.50 |
| Repetition Time (ms) | 7.11 |
| Echo Time (ms) | 2.39 |
| Echo Train Length | 2 |
| Flip Angle (°) | 10 |
| In-plane Resolution (mm) | 1.00 |
| Slice Thickness (mm) | 1.00 |
| Imaging Frequency (Hz) | 63.67 |
| Number Of Averages | 2.00 |
| Percent Sampling (%) | 100 |
| Pixel Bandwidth (Hz) | 405.00 |
| Acquisition Matrix | 256x256 |

#### HOM Cohort T1-w Dixon Water Suppressed SD NMI_c_ Values

**Figure S11.**
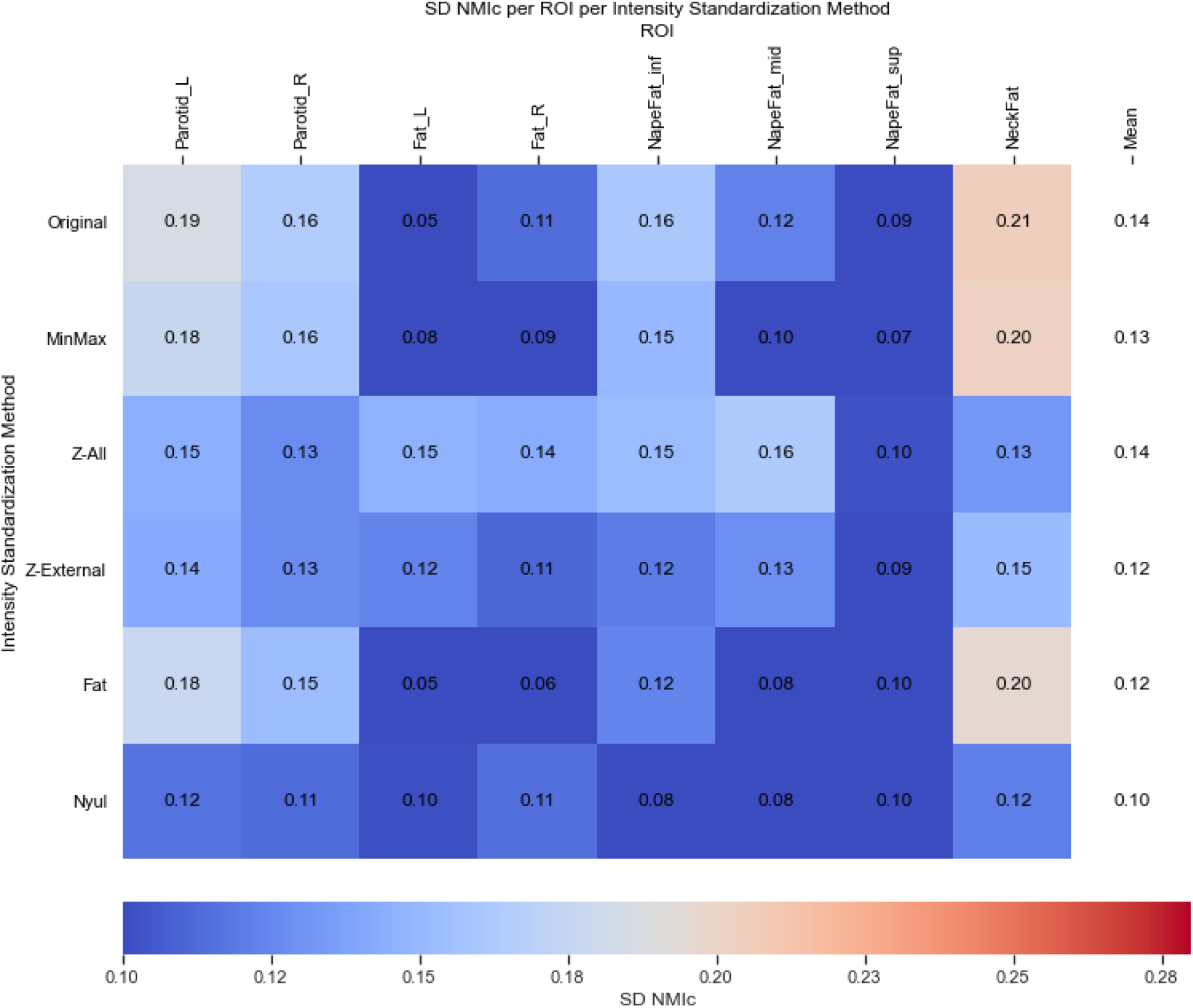
Standard deviation of cohort-level normalized mean intensity (SD NMI_c_) heatmaps per region of interest (ROI) with respect to standardization method for homogeneous (HOM) cohort Dixon T1-weighted Water Suppressed images. The resulting means across all ROIs for each method are shown in the rightmost column of the heatmap. Since water is suppressed in this imaging modality, water-containing structures (muscle, CSF, Cerebellum, etc.) are not visible and were not included in the analysis. Parotids were added as ROIs since they were previously available.

#### HOM Cohort T1-w Dixon Water Suppressed SD NMI_c_ Statistical Comparisons

Friedman test p-value was not significant (p = 0.109) indicating no differences between standardization methods. Therefore, no post-hoc testing was employed.

#### HOM Cohort T1-w Dixon Water Suppressed Histograms

**Figure S12.**
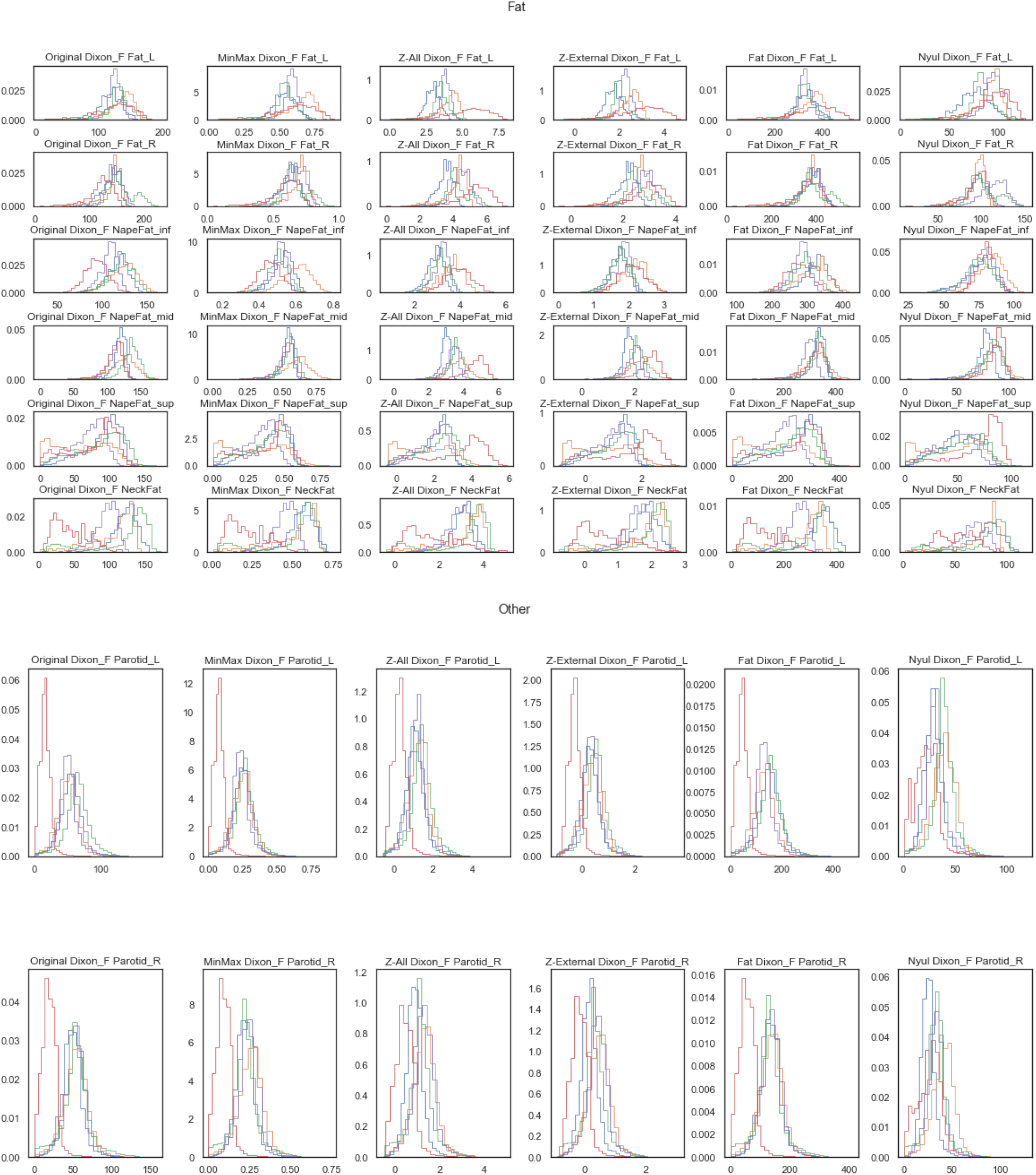
Distributions of region of interest voxel intensities for all patients in the homogeneous (HOM) cohort for Dixon T1-weighted Water Suppressed images. Different colored histograms correspond to different patients in cohort. Columns correspond to methods while rows correspond to regions of interest. First and second sets of plots correspond to Fat and Other categories, respectfully. Image quality was poor for 5^th^ patient (red curve). Interestingly, Nyul method seems to fix the issues so it might be advisable to use this method for future standardization of parotid-related projects dealing with Dixon images of poor quality.

#### HOM Cohort T1-w Dixon Water Suppressed Images

**Figure S13.**
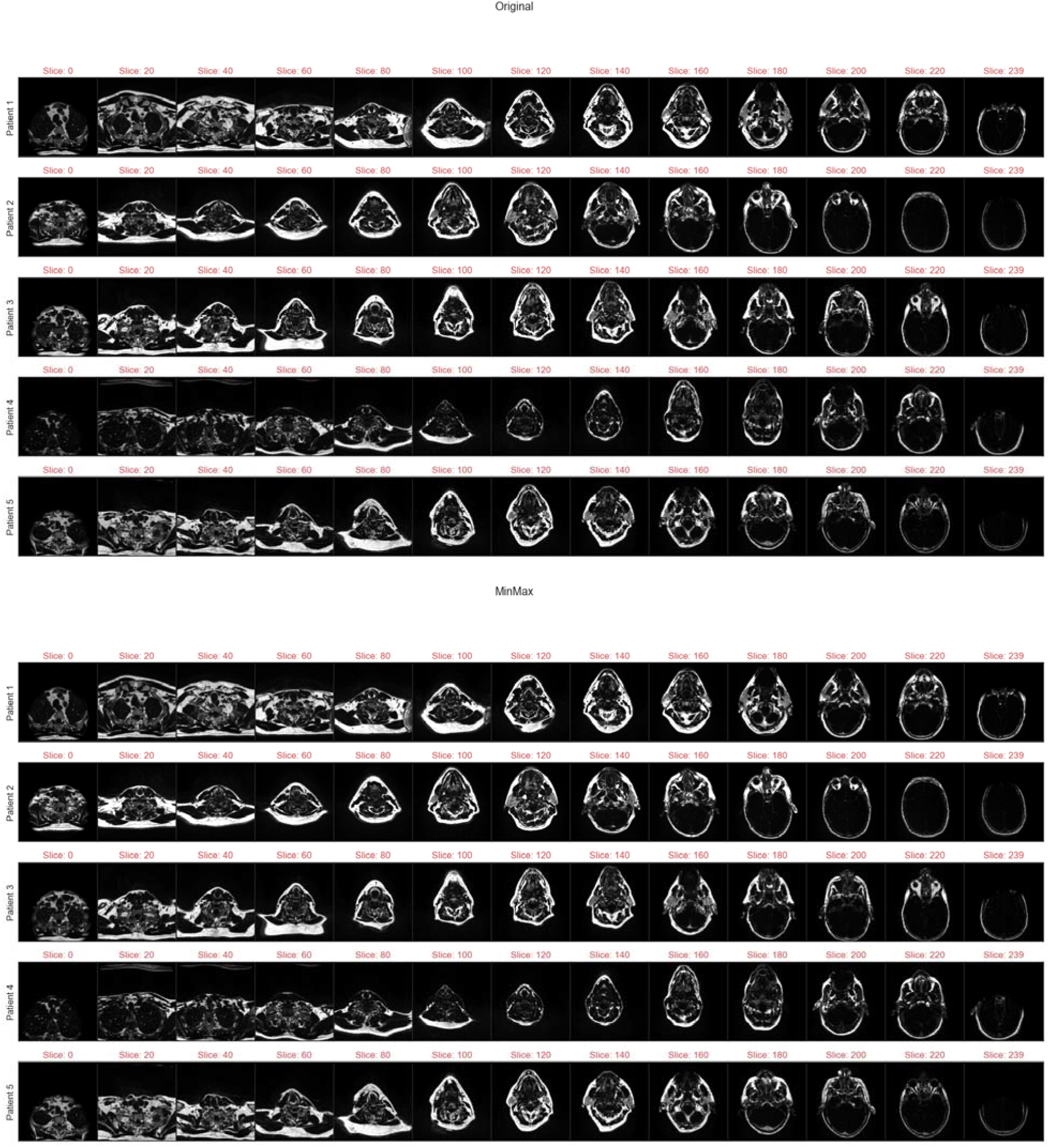
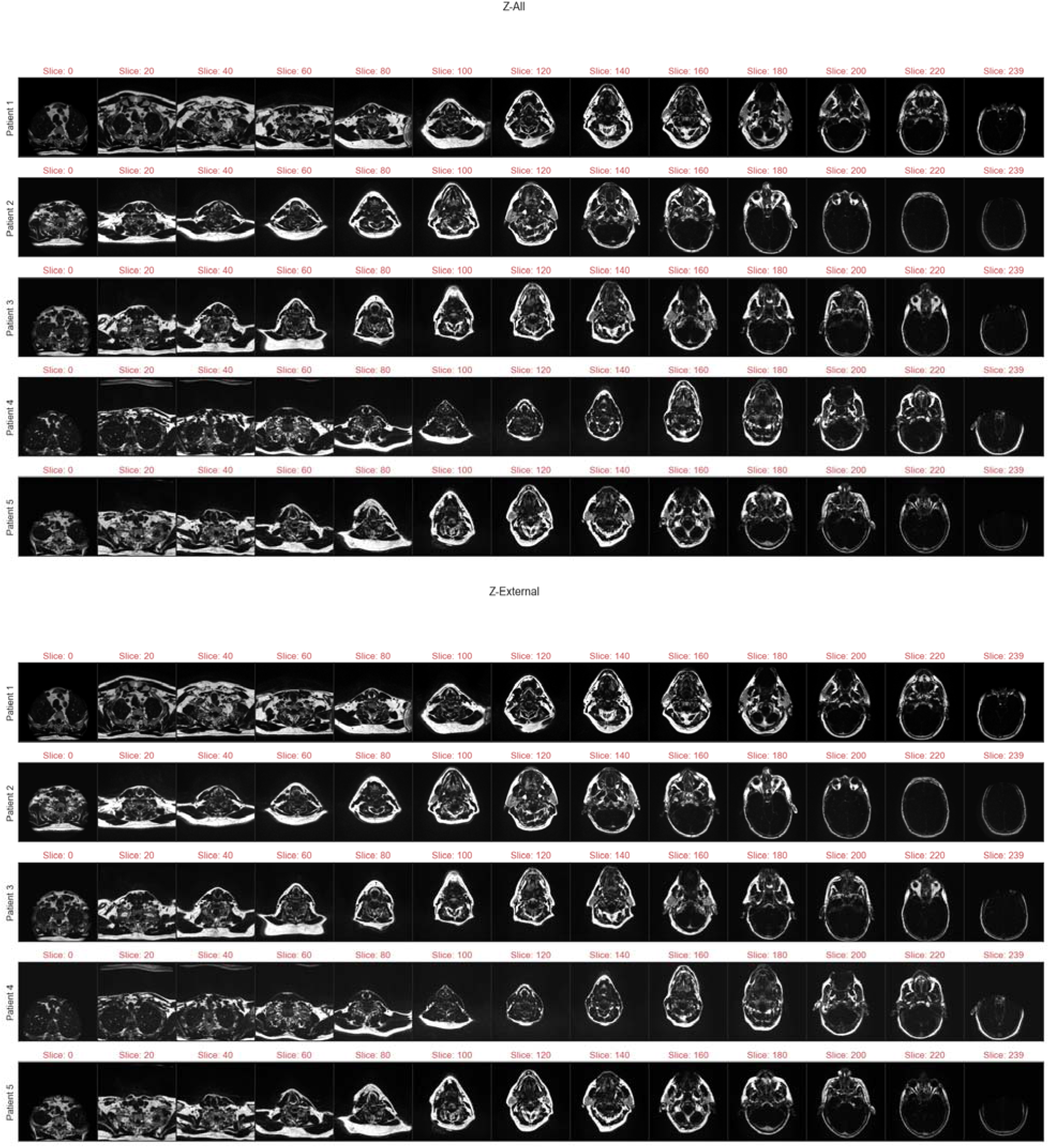
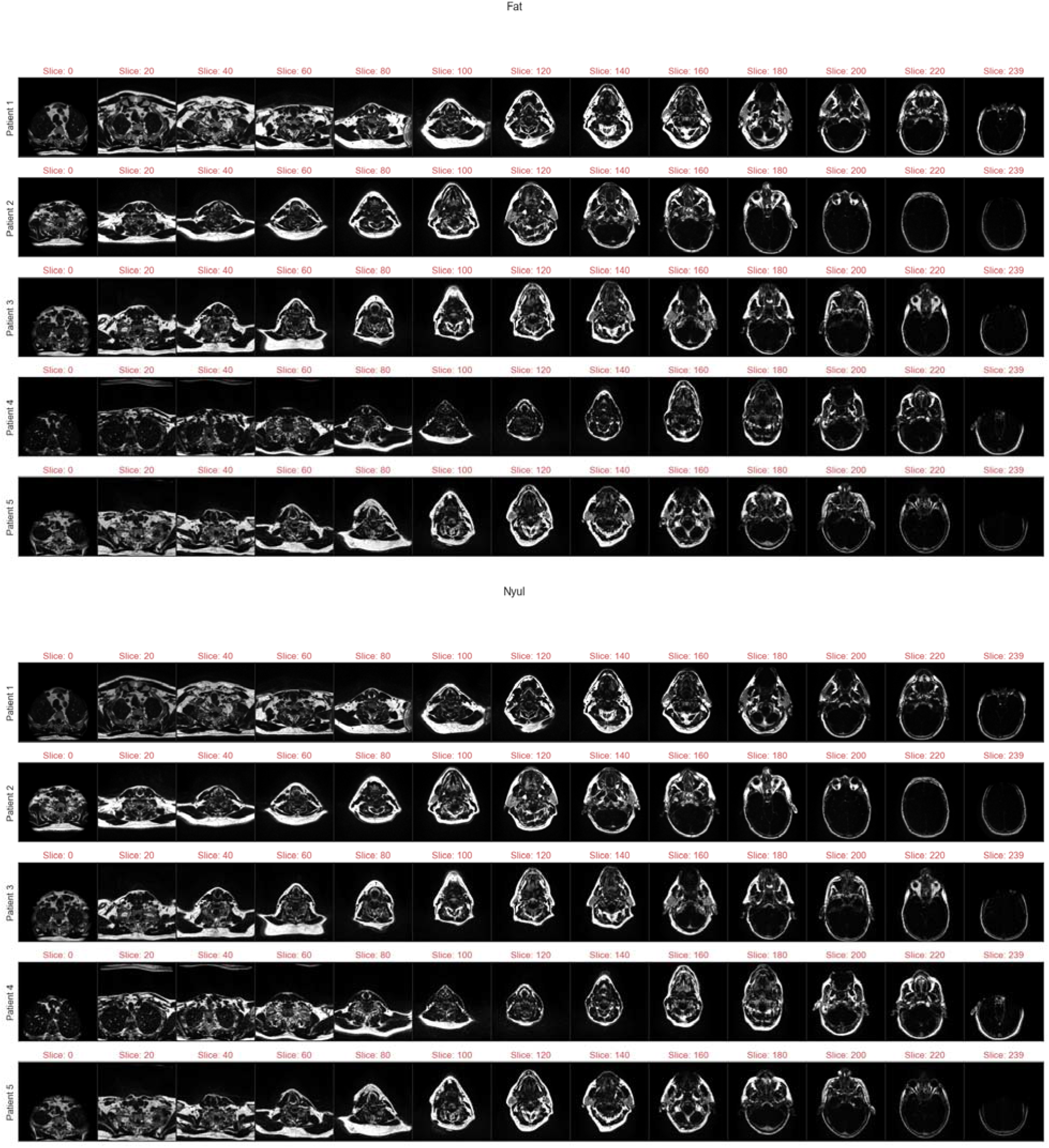
Example Dixon T1-weighted Water Suppressed images for each patient in homogeneous (HOM) cohort. Images are at intervals of 1/13 total image slices to visualize full field of view for each patient. First, second, third, fourth, fifth, and sixth sets of plots correspond to Original, MinMax, Z-All, Z-External, Fat, and Nyul standardization methods, respectively.

### Supplementary Data S5: HOM Cohort CT Data

#### HOM Cohort CT Scanning Parameters

**Table S2.** Homogeneous (HOM) cohort CT scanner characteristics. As opposed to MRI sequences in the HOM cohort, some planning CT images were acquired with different scanners.

| Patient in Cohort | 1 | 2 | 3 | 4 | 5 |
| --- | --- | --- | --- | --- | --- |
| Manufacturer | SIEMENS | SIEMENS | SIEMENS | Philips | Philips |
| Manufacturer Model Name | SOMATOM Definition Edge | SOMATOM Definition Edge | SOMATOM Definition Edge | Brilliance Big Bore | Brilliance Big Bore |
| In-plane Resolution (mm) | 0.97 | 0.97 | 0.97 | 1.17 | 1.03 |
| Slice Thickness (mm) | 2 | 2 | 2 | 3 | 3 |
| Rows | 512 | 512 | 512 | 512 | 512 |
| Columns | 512 | 512 | 512 | 512 | 512 |
| Kilovoltage Peak (kV) | 120 | 120 | 120 | 120 | 120 |
| Convolution Kernel | B40s | B40s | B40s | B | UB |

#### HOM Cohort CT SD NMI_c_ Values

**Figure S14.**
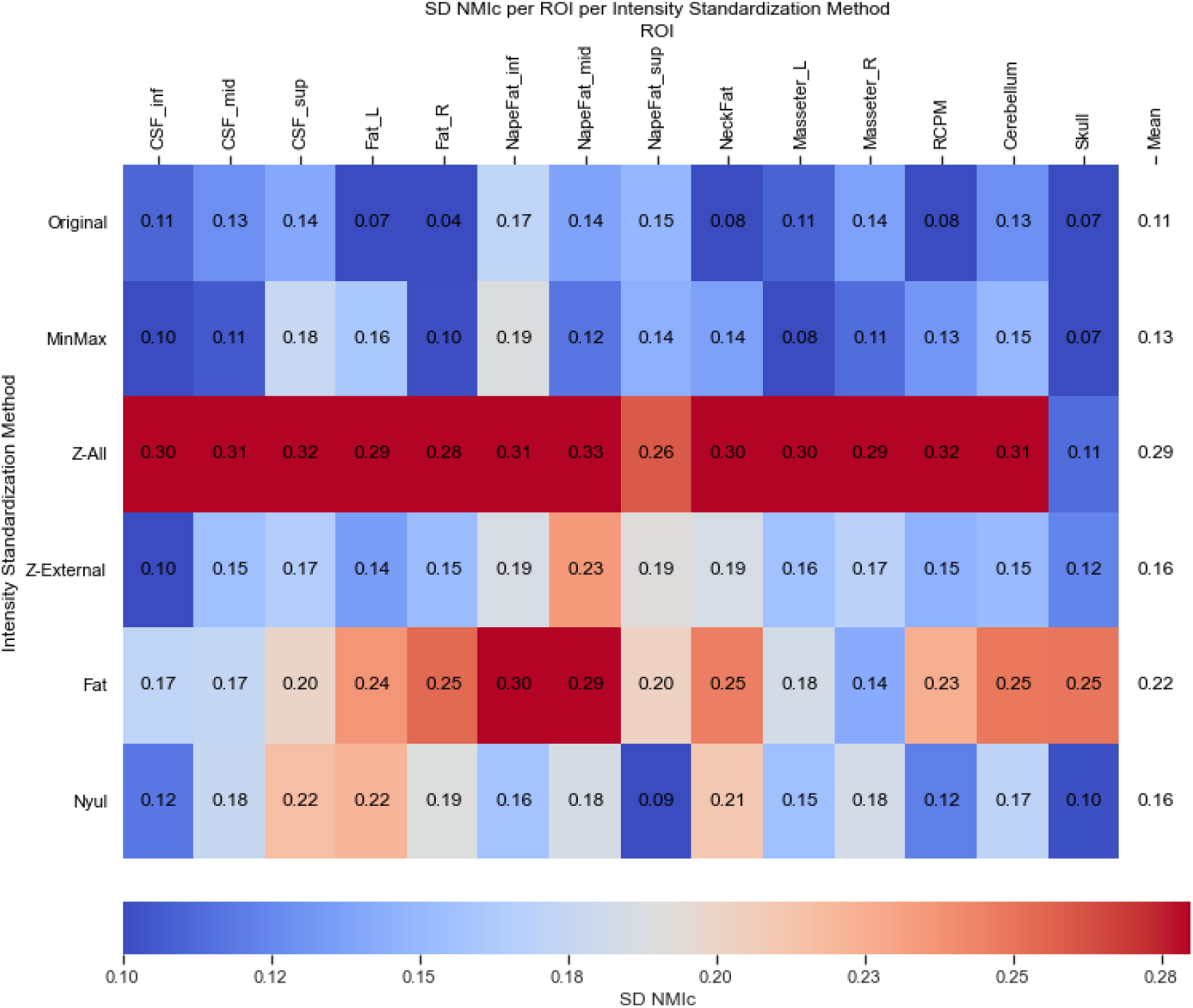
Standard deviation of cohort-level normalized mean intensity (SD NMI_c_) heatmaps per region of interest (ROI) with respect to standardization method for homogeneous (HOM) cohort CT Images. The resulting means across all ROIs for each method are shown in the rightmost column of the heatmap. Means for all standardization methods are worse than unstandardized images. Despite CT being a highly intrinsically standardized imaging modality with a quantitative radiodensity scale, there exists variation across the healthy tissue structures for *Original* images.

#### HOM Cohort CT SD NMI_c_ Statistical Comparisons

**Table S3.** Homogeneous (HOM) cohort CT Friedman test and post-hoc Wilcoxon signed rank test results. * Significant (p<0.05). Standardization normally makes cohort similarity worse for CT, especially a Z-score with all voxels.

| Pairs of standardization methods compared | Wilcoxon signed rank test corrected p-values |
| --- | --- |
| Original vs. MinMax | 1 |
| Original vs. Z-All | 0.001831* |
| Original vs. Z-External | 0.003662* |
| Original vs. Fat | 0.001831* |
| Original vs. Nyul | 0.128174 |
| MinMax vs. Z-all | 0.001831* |
| MinMax vs. Z-External | 0.128174 |
| MinMax vs. Fat | 0.001831* |
| MinMax vs. Nyul | 0.128174 |
| Z-All vs. Z-External | 0.003662* |
| Z-All vs. Fat | 0.128174 |
| Z-All vs. Nyul | 0.001831* |
| Z-External vs. Fat | 0.009155* |
| Z-External vs. Nyul | 1 |
| Fat vs. Nyul | 0.078735 |
| **Friedman test p-value** | **6.294053E-10\*** |

#### HOM Cohort CT Histograms

**Figure S15.**
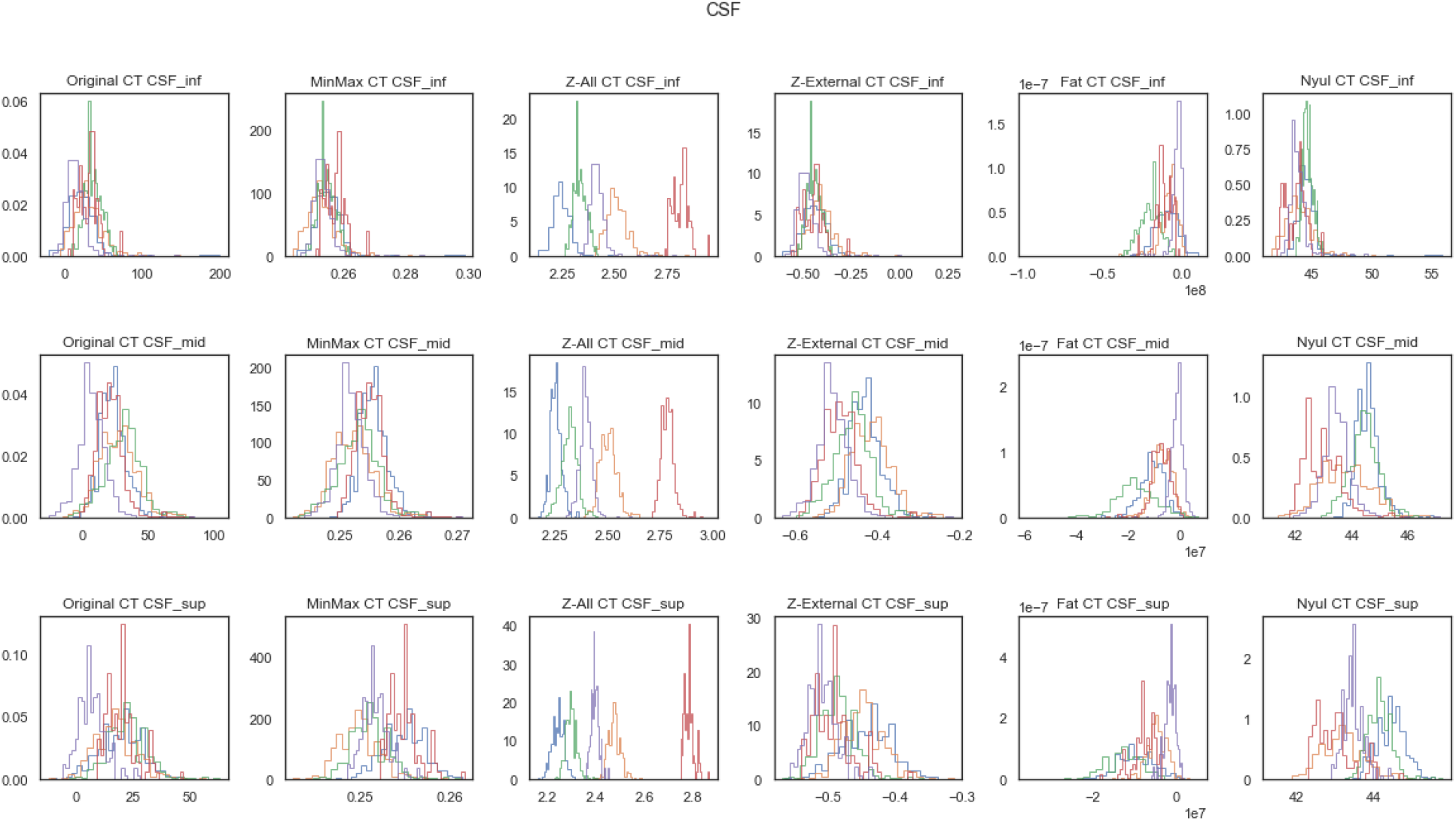
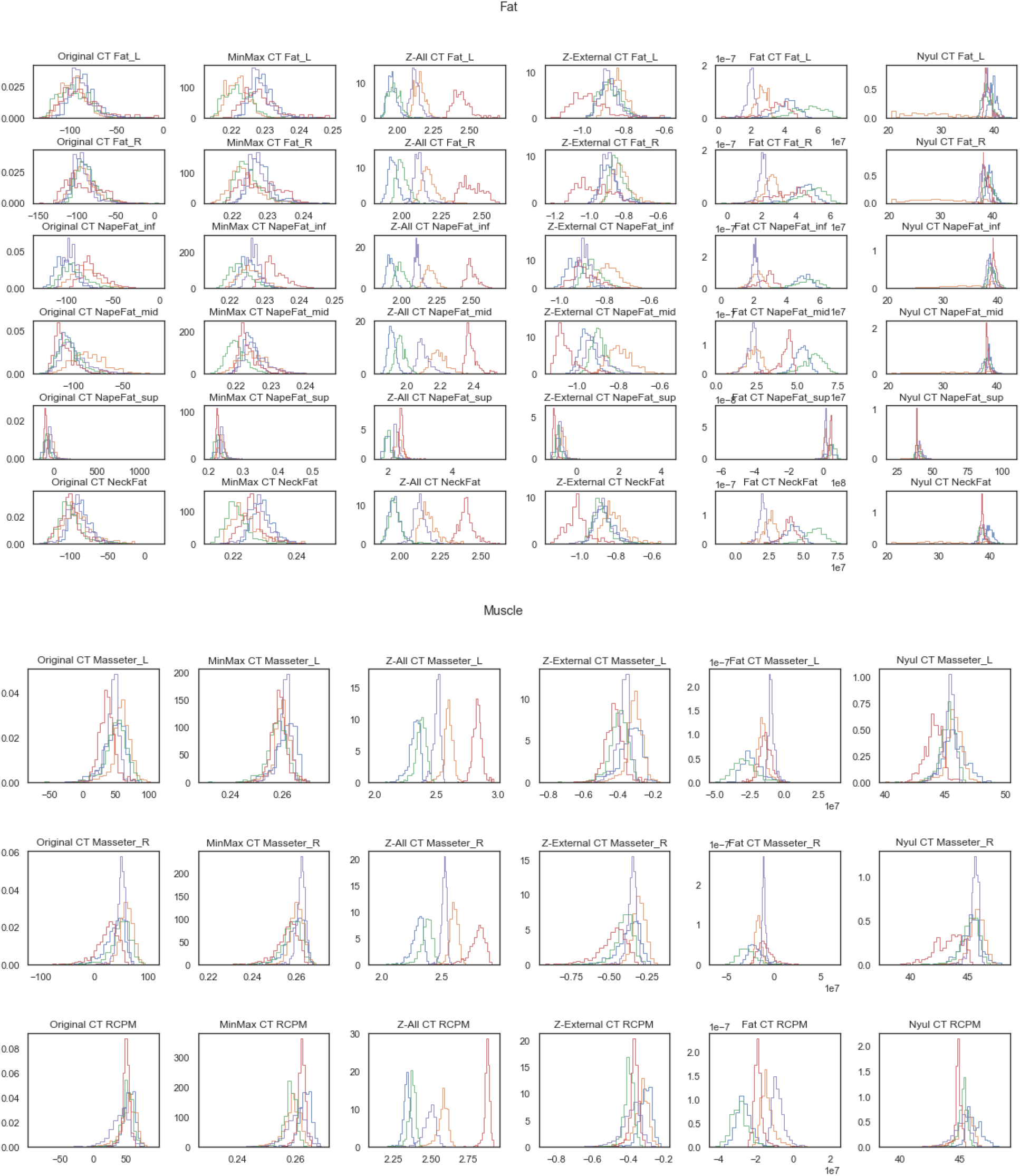
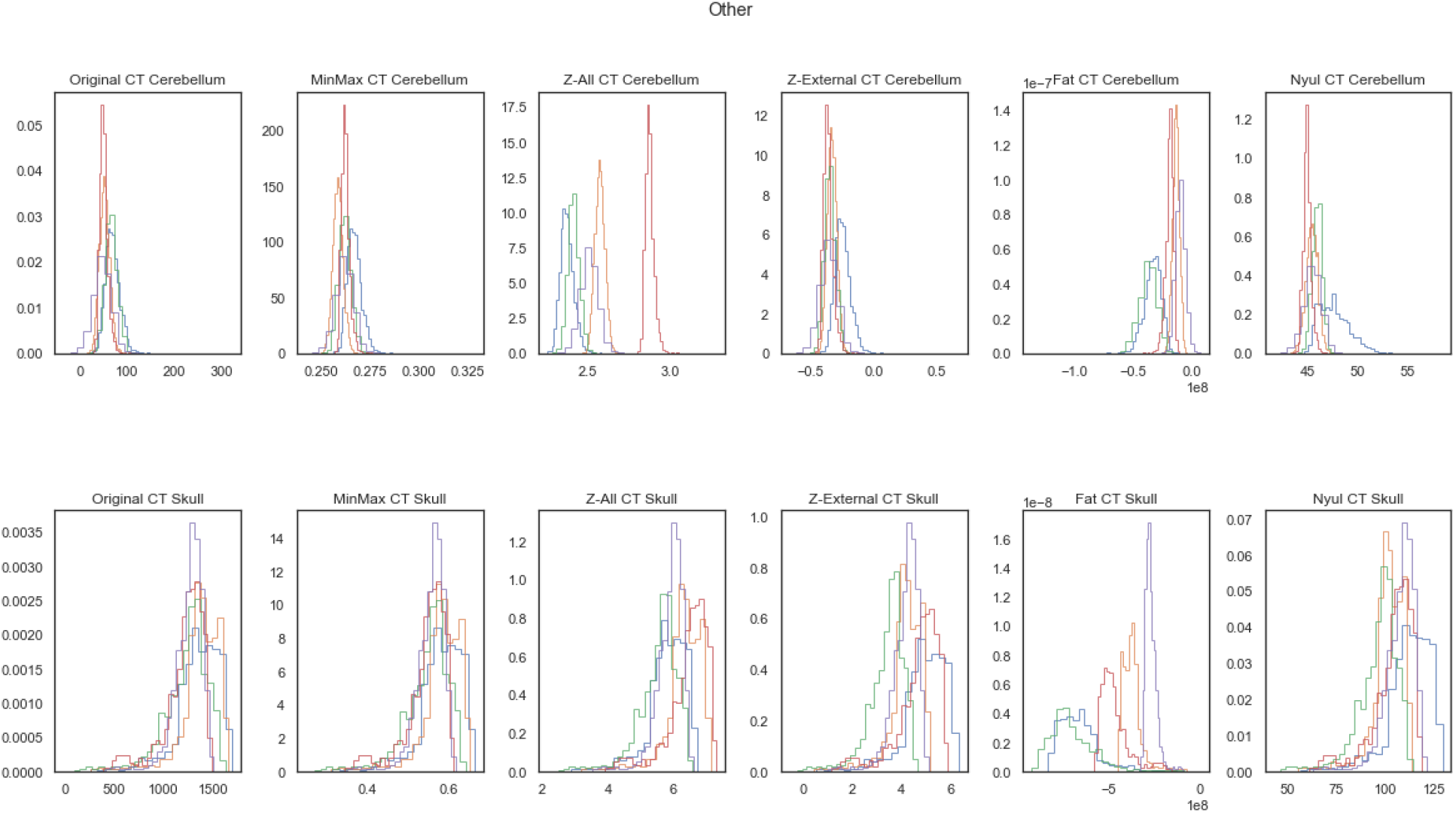
Distributions of region of interest voxel intensities for all patients in the homogeneous (HOM) cohort for CT images. Different colored histograms correspond to different patients in cohort. Columns correspond to methods while rows correspond to regions of interest. First, second, third, and fourth sets of plots correspond to CSF, Fat, Muscle, and Other categories, respectfully.

#### HOM Cohort CT Images

**Figure S16.**
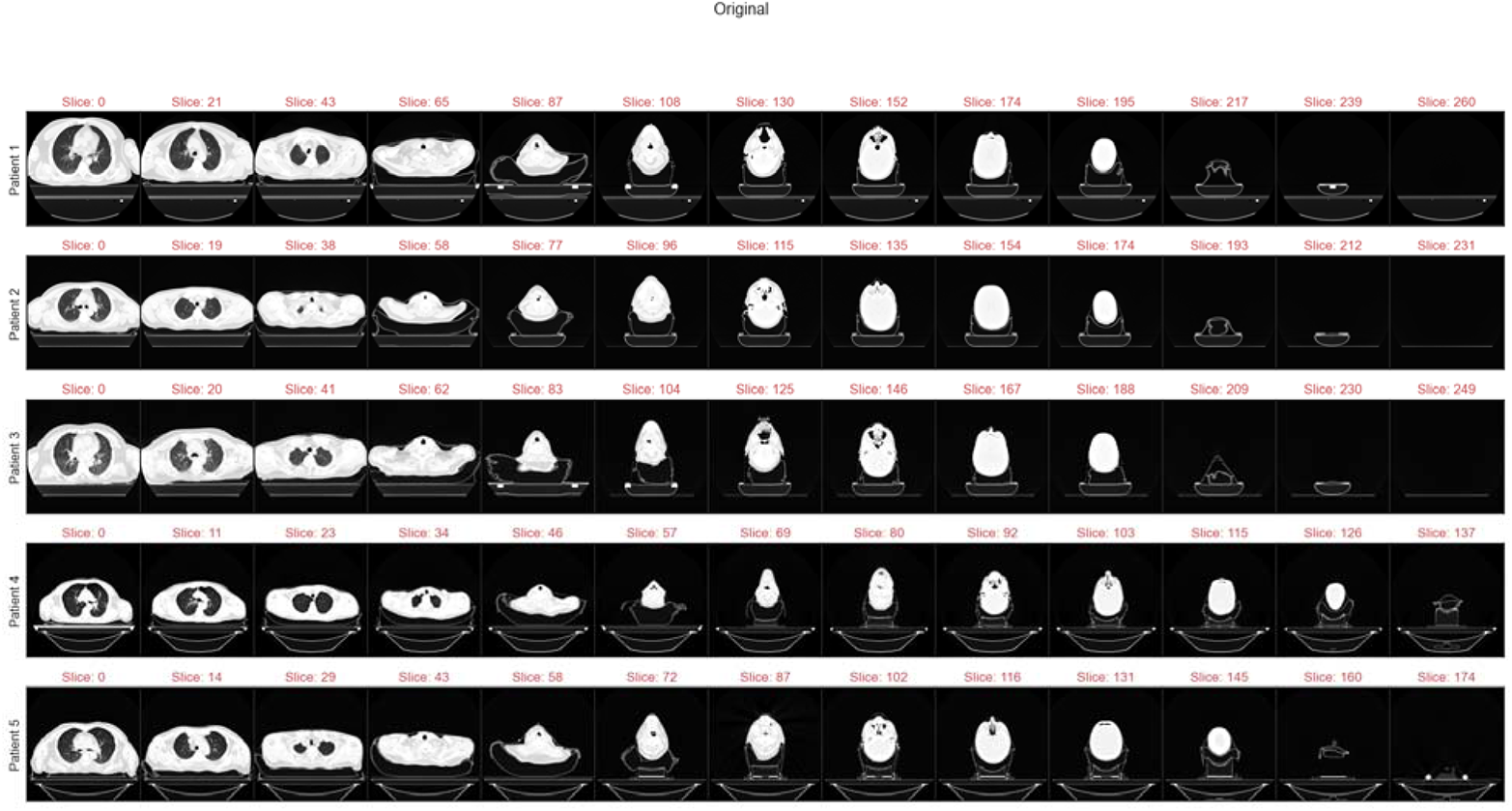
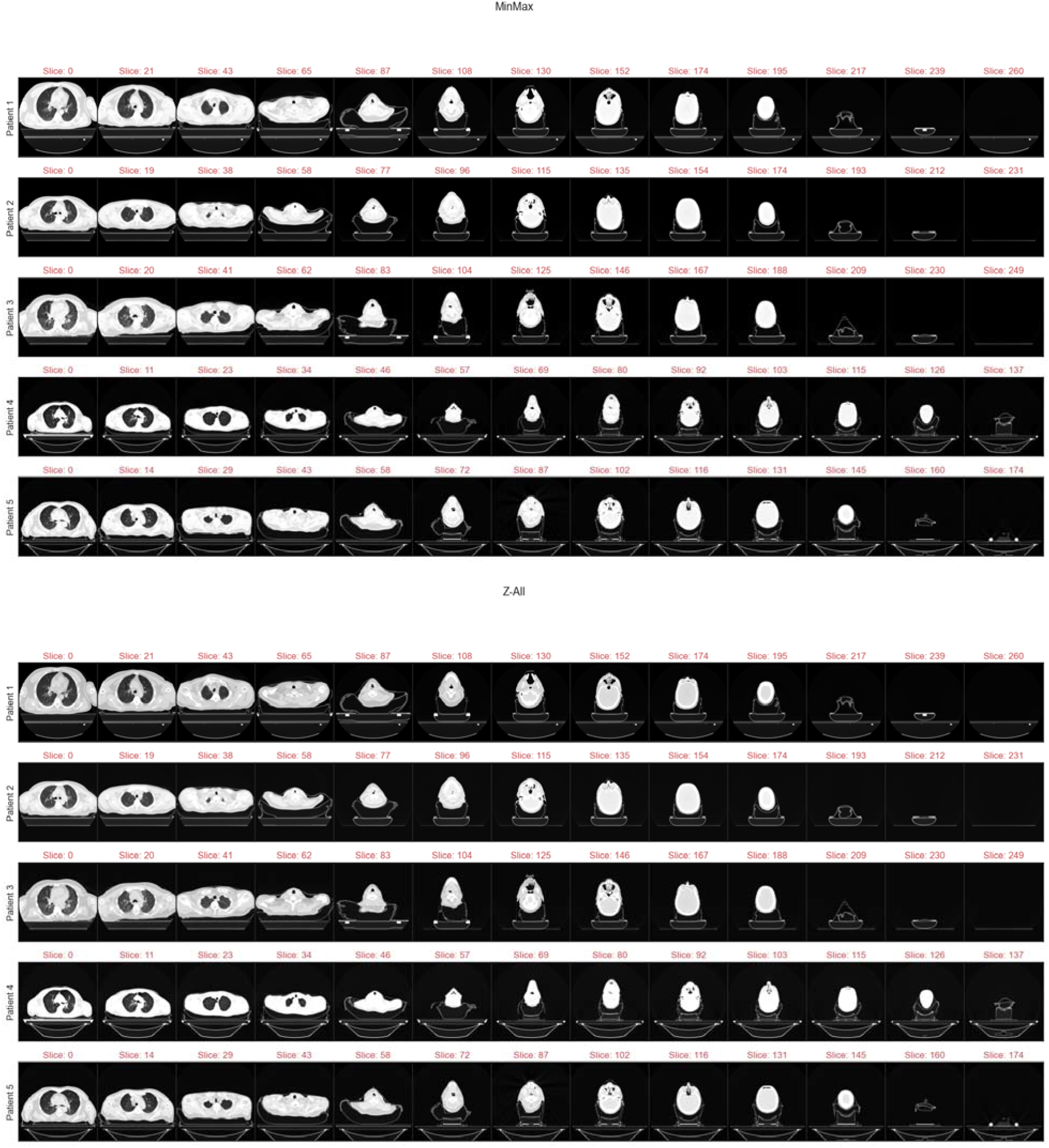
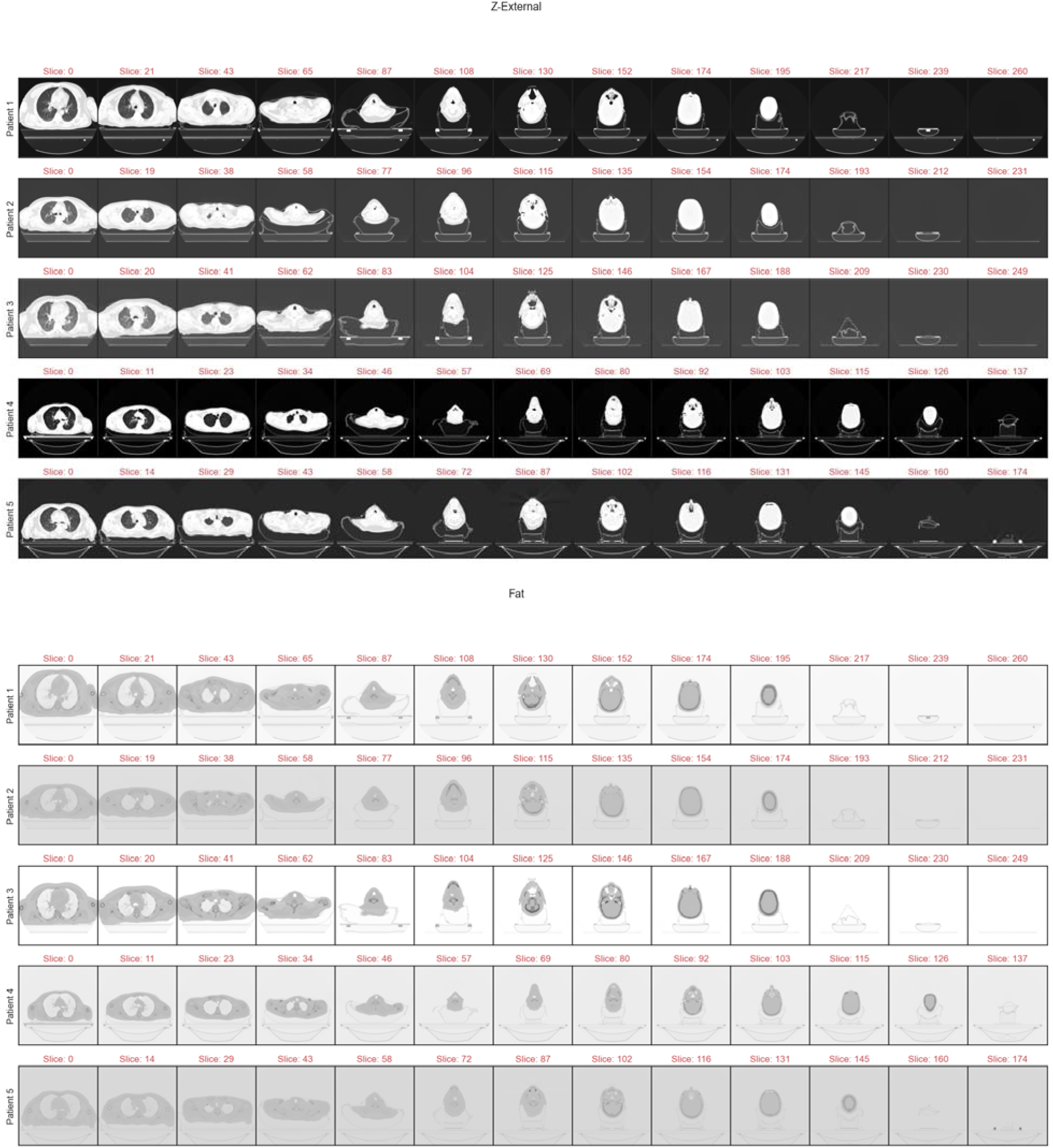
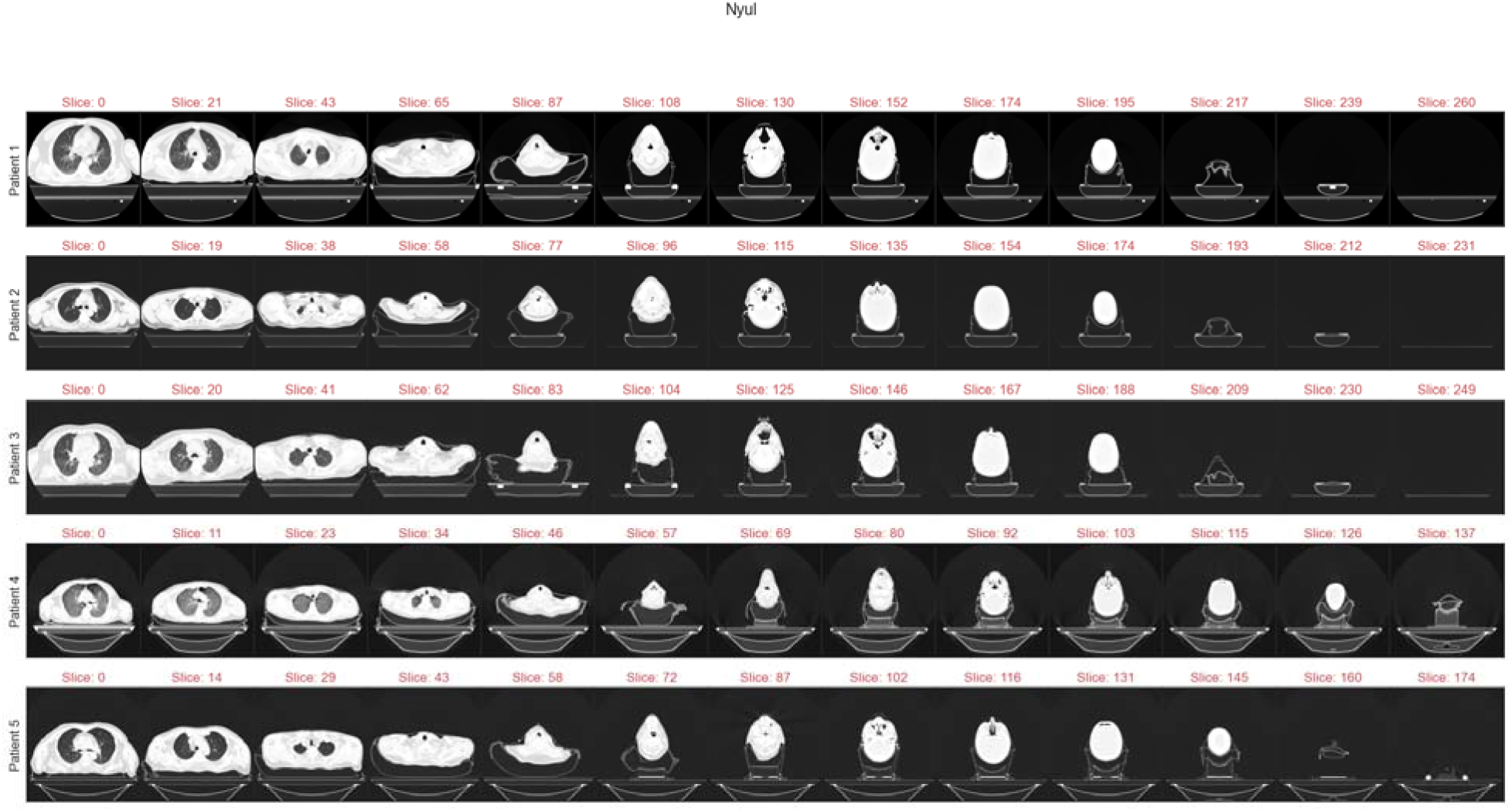
Example CT images for each patient in homogeneous (HOM) cohort. Images are at intervals of 1/13 total image slices to visualize full field of view for each patient. First, second, third, fourth, fifth, and sixth sets of plots correspond to Original, MinMax, Z-All, Z-External, Fat, and Nyul standardization methods, respectively.

### Supplementary Data S6: Radiomic Feature Category Analysis for T2-w MRI

In accordance with the quantitative analysis end goals of MRI intensity standardization, we attempt a cursory examination of radiomic feature classes based on our cohort-level evaluation scheme. Features were extracted for all ROIs using the open-source toolbox PyRadiomics v.3.0 ^3^ for the following feature classes: First Order Statistics, Gray Level Co-occurrence Matrix (GLCM), Gray level Run Length Matrix (GLRLM), Gray Level Size Zone Matrix (GLSZM), and Neighborhood Gray Tone Difference Matrix (NGTDM). Features were extracted with a constant bin number (bin count) of 32, as suggested by previous studies ^4^. Since we are no longer considering a distribution of values but instead a singular value representative of a radiomic feature, SD NMI_c_ is not appropriate as an evaluation metric. Therefore, we instead use the intraclass correlation (ICC) to evaluate the agreement between different standardization methods for a set of ROIs for each feature. We utilize ICC(2,1) where for each standardization method, targets are defined as ROIs, raters are defined as individual patients in a given cohort, and ratings are defined as the numerical value for a given radiomic feature. ICC(2,1) is appropriate since each “subject” is measured by each “rater” and “raters” are considered representative of a larger population, i.e., an arbitrary number of patients ^5^. In principle, a standardization of greater quality should produce more radiomic features that obtain higher ICC values. Here we define ICC quality with the following categorical cutoffs as per previous literature ^5^: <0.5 = Poor, >=0.5 and < 0.75 = Moderate, >= 0.75 and < 0.9 = Good, and >=0.9 = Excellent. We plot the number of features that conform to these categories in Figure S17 for each cohort.

**Figure S17.**
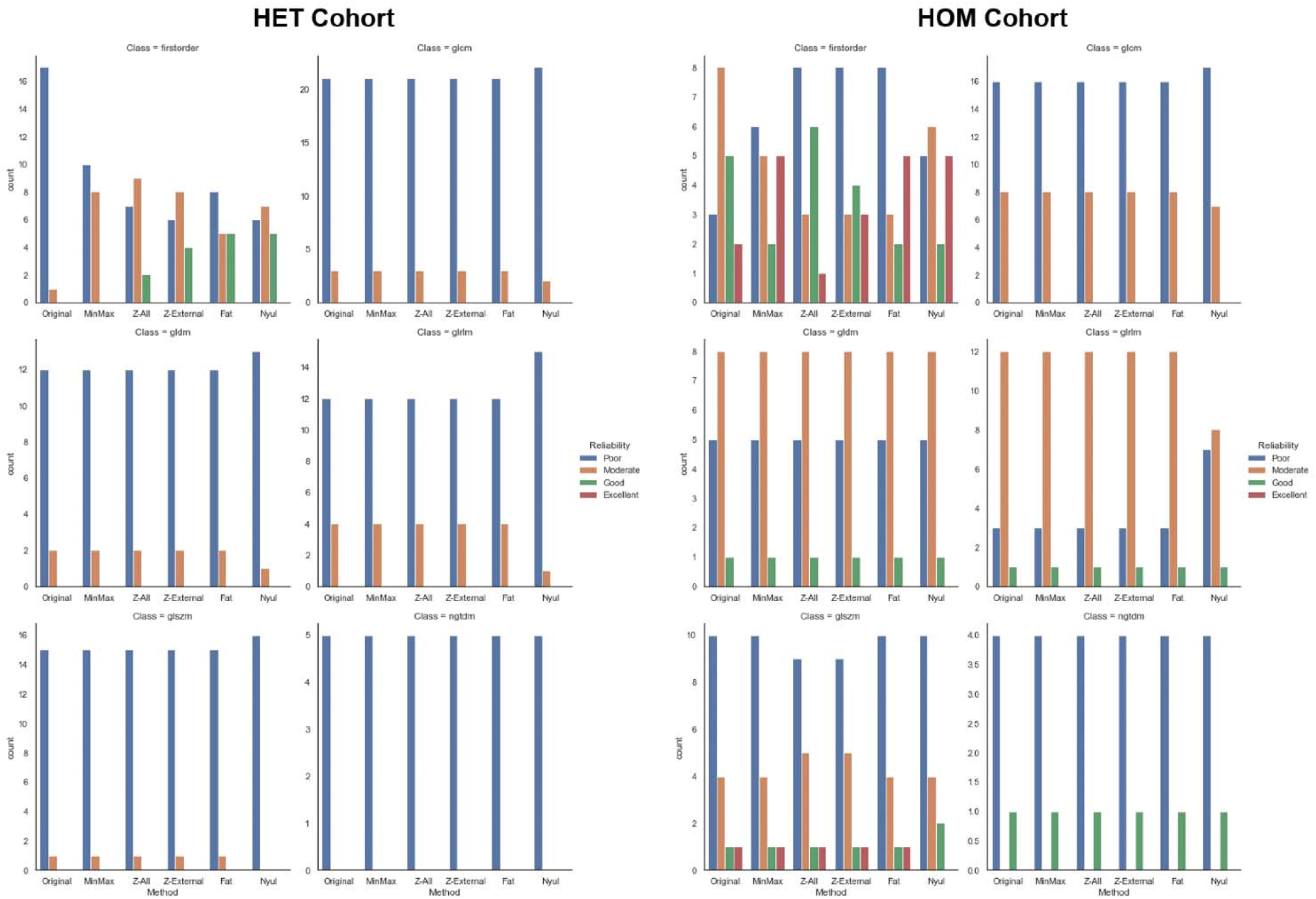
Intraclass correlation (ICC) category (poor, moderate, good, excellent) comparisons between intensity standardization methods per feature category for heterogeneous (HET) cohort (left) and homogeneous (HOM) cohort (right).

We qualitatively observe that for first-order features, similar trends follow our observations for SD NMI_c_. Specifically, in the HET cohort, we observe an increase in similarity with standardization but minimal differences between standardization methods except for *MinMax* which seemingly leads to less reproducible features. Moreover, the HOM cohort shows variation in first-order features between standardization methods, but these differences seemingly balance out in terms of highly reproducible features, suggesting methods are comparable to each other. Trends are less apparent for texture features as most methods lead to indistinguishable ICC stratifications, likely secondary to selecting a constant bin number, as shown in previous studies ^4^. However, some methods, particularly *Nyul*, show slight differences in texture features, leading to increased and decreased reproducibility in different circumstances. Interestingly, regardless of the standardization method, it appears in general the HOM cohort has a greater number of features with higher ICC when compared to the HET cohort, possibly indicating the uniform acquisition parameters playing a larger and more obvious role in radiomic feature reproducibility when compared to simple intensity-based similarity. However, this may also be a byproduct of the choice of gray level discretization ^4,6^. Various investigations in phantoms ^7^ and human subjects ^8^ have shown distinct differences in radiomic features that result from different standardization protocols. Our preliminary results will need to be further investigated to determine what differences lead to higher radiomic reproducibility in images with uniform acquisition parameters.

### Supplementary Data S7: Bias-field in One Sample from HET Cohort

**Figure S18.**
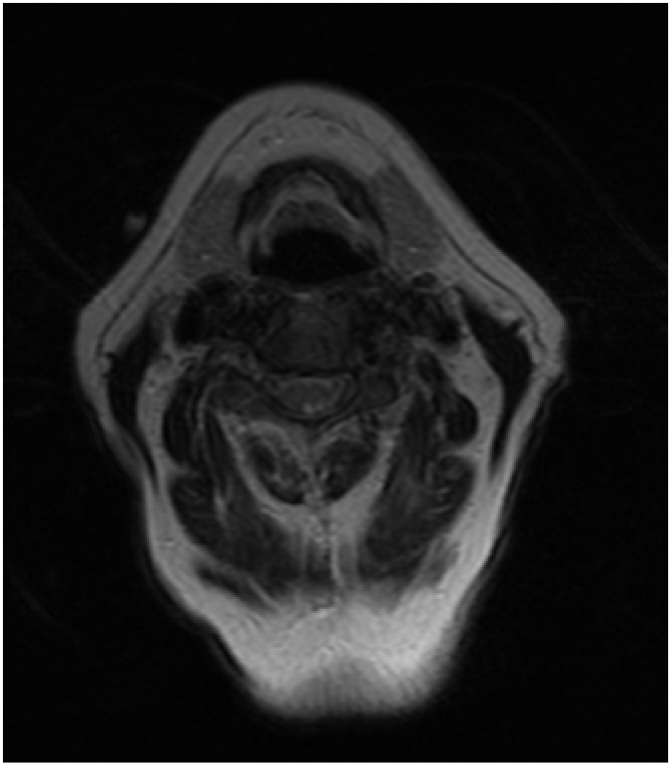
An example of an MRI bias field present in a T2-weighted image from a patient from the heterogeneous (HET) cohort. Bias-field can be visualized in posterior-to-anterior direction.

